# SpinFlowSim: a blood flow simulation framework for histology-informed diffusion MRI microvasculature mapping in cancer

**DOI:** 10.1101/2024.07.15.24310335

**Authors:** Anna Voronova, Athanasios Grigoriou, Kinga Bernatowicz, Sara Simonetti, Garazi Serna, Núria Roson, Manuel Escobar, Maria Vieito, Paolo Nuciforo, Rodrigo Toledo, Elena Garralda, Roser Sala-Llonch, Els Fieremans, Dmitry S. Novikov, Marco Palombo, Raquel Perez-Lopez, Francesco Grussu

## Abstract

Diffusion Magnetic Resonance Imaging (dMRI) sensitises the MRI signal to spin motion. This includes Brownian diffusion, but also flow across intricate networks of capillaries. This effect, the intra-voxel incoherent motion (IVIM), enables microvasculature characterisation with dMRI, through metrics such as the vascular signal fraction *f*_*V*_ or Apparent Diffusion Coefficient (ADC) *D*^*^. The IVIM metrics, while sensitive to perfusion, are in general protocol-dependent, and their interpretation can change depending on the flow regime spins experience during the dMRI measurements (e.g., diffusive vs ballistic), which is in general not known — facts that hamper their clinical utility. Innovative vascular dMRI models are needed to enable the *in vivo* calculation of biologically meaningful markers of capillary flow. These could have relevant applications in cancer, for instance assessing responses to anti-angiogenic therapies targeting tumor vessels. This paper tackles this need by introducing *SpinFlowSim*, an open-source simulator of dMRI signals arising from blood flow within pipe networks. SpinFlowSim, tailored for the laminar flow patterns in capillaries, enables the synthesis of highly-realistic microvascular dMRI signals, given networks reconstructed from histology. We showcase the simulator by generating synthetic signals for 15 networks, reconstructed from liver biopsies, and containing cancerous and non-cancerous tissue. Signals exhibit com-plex, non-mono-exponential behaviours, pointing towards the co-existence of different flow regimes within the same network, and diffusion time dependence. We also demonstrate the potential utility of SpinFlowSim by devising a strategy for microvascular property mapping informed by the synthetic signals, focussing on the quantification of blood velocity distribution moments, and of an *apparent network branching* index. These were estimated *in silico* and *in vivo*, in healthy volunteers and in 13 cancer patients, scanned at 1.5T. In conclusion, realistic flow simulations, as those enabled by SpinFlowSim, may play a key role in the development of the next-generation of dMRI methods for microvascular mapping, with immediate applications in oncology.

## 1 Introduction

In diffusion Magnetic Resonance Imaging (dMRI), water proton motion is encoded in the acquired signals through magnetic field gradients [Kiselev, 2017]. Diffusion encoding provides sensitivity not only to Brownian motion due to pure diffusion, but also to pseudo-diffusion effects arising from the incoherent flow of blood protons through intricate capillary networks [Le Bihan et al., 1986]. Flow through sets of pseudorandomly distributed capillaries leads to magnitude dMRI signal attenuation, a phenomenon known as Intra-Voxel Incoherent Motion (IVIM) effect. IVIM enables the *in vivo* characterisation of microvascular perfusion through dMRI [Le Bihan, 2019], relevant in a variety of diseases, as, for example, in cancer [Fokkinga et al., 2023]. Cancers feature aberrant microvasculature, whose flow patterns can differ considerably from normal tissues [Munn, 2003]. Tumour vasculature is targeted specifically by anti-angiogenic treatments, which are being used in several cancers (e.g., in liver or kidney carcinomas [Jayson et al., 2016]) and tested in combination with therapies such as immune check-point inhibitors, with promising results [Huinen et al., 2021]. The non-invasive assessment of vascular properties through dMRI can equip physicians with new tools for tumour characterisation and longitudinal assessment. It is thereby an active field of research, with studies spanning from malignancy detection to treatment response assessment [Iima et al., 2018, Perucho et al., 2021].

IVIM methods typically rely on disentangling vascular from extra-vascular tissue dMRI signals [Barbieri et al., 2016b, Barbieri et al., 2016a]. Multi-exponential models are routinely used for this purpose, providing metrics such as the vascular signal fraction *f*_^*ν*^_, or the *pseudo-diffusion* (vascular) apparent diffusion coefficient (ADC) *D*^*^. Both *f*_^*ν*^_ and *D*^*^ are useful indices, as they have shown value in cancer assessment [Dappa et al., 2017]. However, these metrics have limitations, since they entangle several, different characteristics of the microvascular scale into a single number, e.g., the product between the means of the blood velocity and capillary length distributions in the diffusive flow regime [Le Bihan and Turner, 1992]. Moreover, they do not account for higher-order cumulants of the diffusion decay (e.g., kurtosis terms proportional to *b*^2^), and their actual numerical value can depend on the acquisition protocol in non-trivial ways [Wu and Zhang, 2019]. In practice, this makes routine IVIM metrics semi-quantitative, surrogate parameters, a fact that, together with their known high variability [Barbieri et al., 2020], hampers their practical clinical deployment.

Recently, the numerical simulation of dMRI signals within histologically-realistic voxel models is being increasingly used to inform parameter estimation [Nilsson et al., 2010, Nguyen et al., 2014, Fieremans and Lee, 2018, Buizza et al., 2021, Morelli et al., 2023]. Simulation-informed approaches increase the realism of signal models, and may thus improve the biological fidelity of dMRI parametric maps [Nedjati-Gilani et al., 2017, Palombo et al., 2019]. However, up to date dMRI simulations have been dominated by Monte Carlo Brownian random walks [Hall and Alexander, 2009, Ginsburger et al., 2019, Rafael-Patino et al., 2020, Lee et al., 2021]. Given that only a few simulation frameworks have focussed on blood flow [Van et al., 2021, Weine et al., 2024], there is an urgent need for new, histologically-meaningful, and reproducible simulation frameworks tailored for dMRI signal arising from blood flow. These could be used to inform novel numerical approaches for non-invasive microvasculature mapping based on dMRI, which could equip oncologists with biologically-meaningful vascular markers in clinical settings. The new dMRI methods could enable the characterisation of capillary flow patterns that are not captured by classical IVIM *f*_^*ν*^_ and *D*^*^, e.g., informing on anisotropic flow patterns, higher-order cumulants or diffusion-time dependence of the vascular signal.

With this article we aim to fill this scientific gap. We present an open-source framework for blood flow simulation within vascular networks, referred to as *SpinFlowSim* from here on, and demonstrate its potential to inform microvasculature property estimation in dMRI. We start by illustrating the computational engine behind SpinFlowSim, based on pipe network theory. Afterwards, we describe the synthesis of dMRI signals arising from flow within realistic vascular networks obtained from histological images of human tumours. Finally, we showcase a potential application of SpinFlowSim, by using the synthetic signals to inform microvasculature property estimation, which is demonstrated *in silico* and *in vivo*, in healthy volunteers and in cancer patients.

## 2 Methods

In this section we illustrate the computational engine upon which SpinFlowSim relies. Afterwards, we present the histological data used to generate realistic vascular networks, and then describe how synthetic dMRI signals were used to inform microvasculature parameter estimation *in silico* and *in vivo*. SpinFlowSim is made freely available at https://github.com/radiomicsgroup/SpinFlowSim.

### 2.1 Simulation framework

In SpinFlowSim we aim to reconstruct the distribution of volumetric flow rate (VFR) across the different segments of an input vascular network. The following characteristics of the vascular network are specified directly by the user:

- a list of capillary segments with their radii;
- the 3D coordinates of the extremities of each segment, referred to as *nodes*;
- the inlet/outlet of the whole network;
- the input VFR *q*_*in*_.

To obtain the VFR distribution, we solve a linear inverse problem, in which the pressure drop Δ*p*_*k,n*_ across each pair of connected nodes (*k, n*) is proportional to the VFR *q*_*k,n*_ between *k* and *n* through a flow resistance coefficient *R*_*k,n*_, via

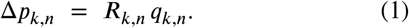

The approach, valid for the laminar flow regime in micro-capillaries, has been recently proposed for capillary flow simulations [Schmid et al., 2015, Van et al., 2021].

To solve for all unknown *q*_*k,n*_ in Eq. 1, we rely on *PySpice* [Salvaire, 2023], a python package for electric circuit analysis, given that our flow problem is formally equivalent to solving a passive electric circuit (electric-hydraulic analogy). Note that in a passive electric circuit, the voltage drop across a resistor is proportional to the product of the electric current through the resistor and the resistance of the element itself, i.e., it is formally equivalent to Eq. 1. In this first demonstration of SpinFlowSim, we compute the resistance between nodes *k* and *n* through a modified Hagen-Poiseuille law, as done in [Blinder et al., 2013]:

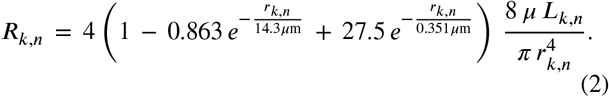

Eq. 2 models the effect of the hematocrit as well as erythrocyte-capillary wall interactions [Pries and Secomb, 2008, Blinder et al., 2013]. Above, *μ* is the dynamic viscosity of pure plasma [Késmárky et al., 2008] (*μ*=1.20 mPa s at 37ºC), *r*_*k,n*_ is the radius of the capillary segment, and *L*_*k,n*_ its length.

After recovering the VFR *q*_*k,n*_ between each pair of connected nodes, in SpinFlowSim we obtain the corresponding mean velocity

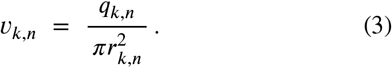

Finally, the 3D trajectory **p**_*w*_(*t*) of the generic *w*-th blood spin is synthesised by integrating the discrete-time system

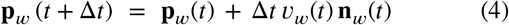

given an initial position **p**_*w*_(0) = **p**_*w*,0_. In Eq. 4, Δ*t* is the temporal resolution of the simulation, while *ν*_*w*_(*t*) and **n**_*w*_(*t*) are the instantaneous velocity vector magnitude and direction experienced by the spin at time *t*. Spins’ initial positions **p**_*w*,0_ are seeded across the whole network, with uniform spin density in each segment. The numbers of spins assigned to each segment is proportional to its volume [Van et al., 2021]. During the integration of Eq. 4, spins reaching the termination of a capillary are assigned at random to one of the emanating branches. The probability of a spin being assigned to a specific branch is proportional to the VFR through that branch [Van et al., 2021]. More formally, once a flowing spin reaches the *k*-th node, the probability of it continuing its trajectory in the *k* → *n* branch emanating from *k* is 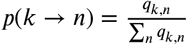. Moreover, spins reaching the network outlet continue flowing through a shifted copy of the vascular network, whose inlet position coincides exactly with the outlet itself. This ensures that no spins are lost during the simulation (periodic boundary condition). SpinFlowSim supports the visualisation of spin trajectories as a video, in order to facilitate the visual inspection of the simulation output.

Once the trajectories for *W* spins have been generated, we synthesise a complex-valued dMRI signal *s* for any input gradient wave form **G**(*t*) as [Fieremans and Lee, 2018]

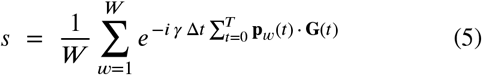

given the requested total simulation duration *T*.

### 2.2 Vascular networks

We deployed SpinFlowSim on vascular networks reconstructed from 2D histological images. These consisted of biopsies obtained in patients suffering from advanced tumours and participating in an ongoing imaging study at the Vall d’Hebron Institute of Oncology (Barcelona).

The biopsied tissue, taken from liver tumours, was processed and stained. Digitised images of the stained tissue were acquired on a Hamamatsu C9600-12 optical slide scanner (resolution: 0.454 *μm*). For this study, we used 11 histological images, obtained from 11 patients. For each patient, we had access to either a routine hematoxylin-eosin (HE) stain (n=9) or a CD31 stain (n=2).

We drew a total of 15 2D networks. We drew networks manually, by tracing visible capillaries in non-cancerous liver parenchyma or in cancerous regions-of-interest (ROIs). Networks were drawn on approximately square ROIs, of sizes ranging from 250 to 550 *μm* per side. Networks were made of interconnected segments, with curved capillaries approximated by a piece-wise series of straight pipes. A characteristic radius was assigned to each segment by averaging three radius measurements, performed on at the inlet, middle point, and outlet level. For each network, we computed an approximated network size as the maximum euclidean distance between any pair of nodes. We also computed the mean and standard deviation of the capillary radii and lengths (*r* and *L*).

We generated 100 VFR distribution realisations by changing randomly the position of the network inlet/outlet 10 times, and varying the input VFR *q*_*in*_ for each inlet/outlet pair (10 uniformly-spaced *q*_*in*_ values in [1.5*⋅*10^−4^; 5.5*⋅*10^−3^] mm^3^/s), to cover plausible blood capillary velocities [Ivanov et al., 1981]. The total duration and the temporal resolution of the simulations were *T* = 100 *ms* and Δ*t* = 0.01 *ms*. We characterised each realisation by computing the mean (*ν*_*m*_) and standard deviation (*ν*_*s*_) of the velocity distribution across capillary segments, as well an Apparent Network Branching (*ANB* index). *ANB* measures the average number of segments spins travel through during the simulation. Spearman’s correlation coefficients among all possible pairs of mean *ν*_*m*_, *ν*_*s*_, *ANB, r* and *L* were computed.

Finally, we synthesised illustrative dMRI signals for routine pulsed-gradient spin echo (PGSE) sequences. We probed b-values in the range [0; 150] s/mm^2^, and varied the gradient separation Δ in Δ = {30, 50, 70} ms, while fixing the gradient duration to *δ* = 20 ms. Signals were generated for two orthogonal directions within the plane containing the 2D networks, and their magnitude averaged. Signals were characterised through the computation of the vascular ADC *D*^*^ via linear fitting (minimum/maximum *b* of 0/100 s/mm^2^, with 10 s/mm^2^ increments). We investigated whether microvascular properties are encoded in the diffusion-weighted (DW) signal by scattering *D*^*^ against each of *ν*_*m*_, *ν*_*s*_ and *ANB* in turn, at fixed gradient separation Δ. Linear fitting between *D*^*^ and each microvascular property was performed and Spearman’s correlation coefficients were computed, in order to corroborate qualitative trends on scatter plots.

### 2.3 Microvascular property estimation from dMRI

We also investigated whether the synthetic signals generated with SpinFlowSim can be used to inform microvascular parameter estimation in dMRI. We hypothesised that, for a given dMRI protocol, large dictionaries of synthetic, noise-free signal arrays **S** = {**s**_1_, …, **s**_*M*_ }, coupled with their corresponding vascular parameter arrays **P** = {**p**_1_, …, **p**_*M*_ }, can be used to find practical numerical implementations of the forward signal model **P** ↦ **S**(**P**). Numerical implementations of this type could be easily incorporated in standard non-linear least square (NNLS) fitting, used routinely in dMRI, thus avoiding the need for approximated analytical signal expressions.

In the following sections, we will describe *in silico* analyses performed to investigate the feasibility of simulation-informed fitting. We will then describe experiments performed to demonstrate the approach *in vivo*, based on the acquisition of dMRI scans in healthy volunteers and cancer patients at 1.5T.

#### 2.3.1 *In silico* estimation

We used SpinFlowSim to synthesise signals for 3 realistic dMRI protocols, and then analysed such signals to test whether it is possible to estimate *ν*_*m*_, *ν*_*s*_ and *ANB* from noisy measurements. One of the protocols represents a rich, comprehensive PGSE acquisition, encompassing several b-values in a measurement regime with high sensitivity to IVIM effects (i.e., *b* smaller than approximately 100 s/mm^2^ [Le Bihan, 2019]), as well as multiple diffusion times. A second protocol is instead a shorter subset of the rich protocol. Finally, the third protocol matches a DW twice-refocussed spin echo (TRSE) used for *in vivo* imaging. Signals were generated for two orthogonal directions within the plane containing the 2D networks, and their magnitude averaged. In summary, the protocols were:

- a rich PGSE protocol, referred to as “richPGSE”. It consisted of a total of 99 measurements, consisting of 9 *b* = 0 and 10 non-zero b-values *b* = {10, 20, 30, 40, 50, 60, 70, 80, 90, 100} s/mm^2^, each acquired for 9 unique diffusion times, corresponding to (*δ*, Δ) = {10, 20, 30} ms × {30, 50, 70} ms.
- A second PGSE protocol, referred to simply as “PGSE”. It is a subset of the former, and describes a more realistic acquisition that could be implemented under time pressure. It encompassed 3 *b* = 0 and 6 diffusion-weighted (DW) measurements, namely *b* = {50, 100} for 3 different diffusion times. The gradient duration *δ* was fixed to 20 ms, while the 3 diffusion times were achieved by varying Δ as Δ = {30, 50, 70} ms.
- a DW twice-refocussed spin echo (TRSE) protocol, referred to simply as “TRSE”. It matches the protocol implemented on a 1.5T Siemens Avanto system *in vivo* (see section 2.3.2 below). It consisted of 3 non-DW and 6 DW measurements. These were *b* = {50, 100}, acquired for 3 diffusion times. The gradient duration of the 4 gradient lobes (Supplementary Fig. 1) for the 3 diffusion times were: *δ*_1_ = {8.9, 13.2, 18.9} ms, *δ*_2_ = {17.6, 19.3, 21.0} ms, *δ*_3_ = {20.4, 24.8, 30.5} ms, *δ*_4_ = {6.0, 7.7, 9.5} ms. The separation between the onset of the gradient lobes (Supplementary Fig. 1) were instead: Δ_1,2_ = {17.4, 21.7, 27.5} ms, Δ_1,4_ = {63.9, 74.2, 87.5} ms.

Briefly, we performed a leave-one-out experiment. For each vascular network in turn, we used noise-free signals from 14 out of 15 substrates to learn the forward signal model (*ν*_*m*_,*ν*_*s*_, *ANB*) ↦ *s*(*ν*_*m*_, *ν*_*s*_, *ANB*), which we then used for estimating *ν*_*m*_, *ν*_*s*_ and *ANB* on noisy signals for the remaining 15-th network (signal-to-noise ratio (SNR) at *b* = 0 of 20). The forward signal model (*ν*_*m*_, *ν*_*s*_, *ANB*) ↦ *s*(*ν*_*m*_, *ν*_*s*_, *ANB*) was learnt by interpolating the set of paired examples signals/vascular parameters with a radial basis function (RBF) regressor, so that fitting could be performed by embedding *s*(*ν*_*m*_, *ν*_*s*_, *ANB*) into standard maximum-likelihood NNLS routines [Panagiotaki et al., 2012]. Fitting was performed with the freely-available *mri2micro*_*dictml*.*py* tool, part of the *bodymri-tools* python repository (https://github.com/fragrussu/bodymritools). To characterise fitting performance, we generated scatter plots between ground truth and estimated *ν*_*m*_, *ν*_*s*_ and *ANB*, and computed corresponding Spearman’s correlation coefficients.

#### 2.3.2 *In vivo* estimation

We also investigated the feasibility of using synthetic signals from SpinFlowSim to inform microvascular property estimation *in vivo*, on both healthy volunteers and cancer patients. All participants were scanned after providing informed written consent, in imaging sessions approved by the Clinical Research Ethics Committee (CEIm) of the Vall d’Hebron University Hospital of Barcelona, Spain (codes: PR(AG)29/2020 and PR(IDI)109/2022)).

##### Healthy volunteers: data and analysis

We scanned two healthy volunteers in their thirties on a 1.5T Siemens Avanto system. The acquisition included routine anatomical imaging and a DW TRSE Echo Planar Imaging (EPI) scan, with salient parameters: resolution of 1.9 × 1.9 × mm^2^; slice thickness of 6 mm; *b* = {0, 50, 100, 400, 900, 1200, 1600} s/mm^2^, with each *b* acquired at 3 different diffusion times, with the same diffusion times as the “TRSE” protocol used simulations (see section 2.3.1 above); TE = {93, 105, 120} ms for the short, intermediate, and long diffusion time; TR = 7900 ms; trace DW imaging; NEX = 2; GRAPPA = 2; 6/8 Partial Fourier imaging; BW = 1430 Hz/pixel; acquisition of a *b* = 0 image at the shortest TE with reversed phase encoding.

We denoised scans with MP-PCA [Veraart et al., 2016], mitigated Gibbs ringing [Kellner et al., 2016] and corrected for motion and EPI distortions [Andersson et al., 2003]. Subsequently, we normalised the signal acquired at each TE to the *b* = 0 signal level at the same TE, and then estimated the vascular signal *S*_*V*_ for *b* ≤ 100 s/mm^2^ in each voxel [Gurney-Champion et al., 2018, Wang et al., 2021], as

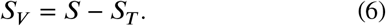

Above, *S* is the measured signal and *S*_*T*_ is an estimate of the extra-vascular tissue signal. *S*_*T*_ was computed by extrapolating to *b* ≤ 100 s/mm^2^ an ADC fit 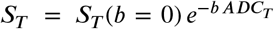 performed on signal measurements at *b* > 100 s/mm^2^.

Afterwards, we fitted (*ν*_*m*_, *ν*_*s*_, *ANB*) voxel-by-voxel, using the same fitting procedure employed in *in silico* experiments above, but learning the forward model **P** ↦ **S**(**P**) on all 1500 synthetic signals from all vascular networks. For reference, we also computed more standard IVIM metrics *f*_*V*_ and *D*^*^, by fitting 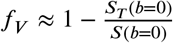 to the vascular signal estimated at the shortest TE, with 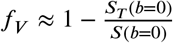. Mean and standard er-rors of *ν*_*m*_, *ν*_*s*_, *ANB, f*_*V*_ and *D*^*^ within manually drawn ROIs were computed. The ROIs were placed in the liver, spleen, as well as medulla and cortex of a kidney.

##### Cancer patients: data and analysis

Finally, we tested our simulation-informed parameter estimation on dMRI scans of 13 patients suffering from advanced solid tumours (7 females, 5 males; approximate age range 30-85 y.o.), who participated in an ongoing imaging study at the Vall d’Hebron Institute of Oncology (Barcelona, Spain). Patients were scanned on the same 1.5T Siemens Avanto system used to acquire data on healthy volunteers, and according to the same imaging protocol. dMRI scans underwent the same processing as described above, obtaining voxel-wise maps of *ν*_*m*_, *ν*_*s*_, *ANB, f*_*V*_ and *D*^*^. Mean and standard deviation of such metrics within tumours were obtained, with tumours manually segmented by an expert radiologist (R.P.L).

## 3 Results

### 3.1 Vascular networks

Fig. 2 illustrates the 15 vascular networks generated in this study from liver tumour biopsies. Out of the total, 3 were segmented on non-cancerous liver parenchyma, while the remaining 12 on cancerous tissue. The 3 non-cancerous networks were drawn on liver tissue found on the histological slide, adjacent to tumour tissue (n=2 melanoma metastases; n=1 ovarian cancer metastasis). The 12 networks drawn on cancerous tissue came from primary liver hepatocellular carcinoma (HCC, n=5), or from liver metastases of colorectal cancer (CRC, n=5), endometrial cancer (n=1), and melanoma (n=1).

**Figure 1:**
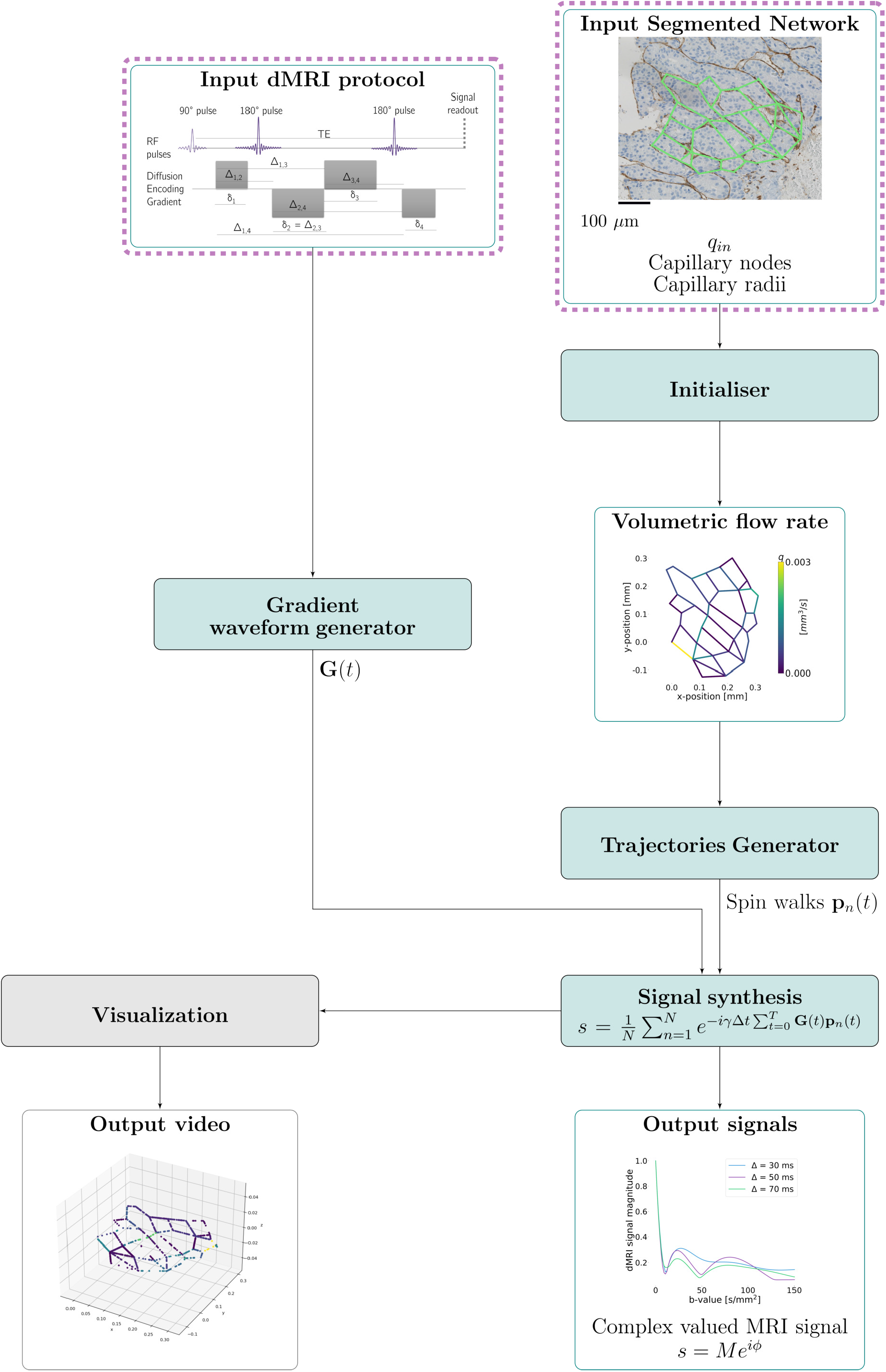
Outline of the proposed SpinFlowSim framework. The dashed red box indicates user-provided input information. An illustrative example of a network segmented on a biopsy with resolved volumetric flow rates for an input flow *q*_*in*_ = 3.1 *⋅* 10^−3^ mm^3^/s and synthesised signals are shown.

**Figure 2:**
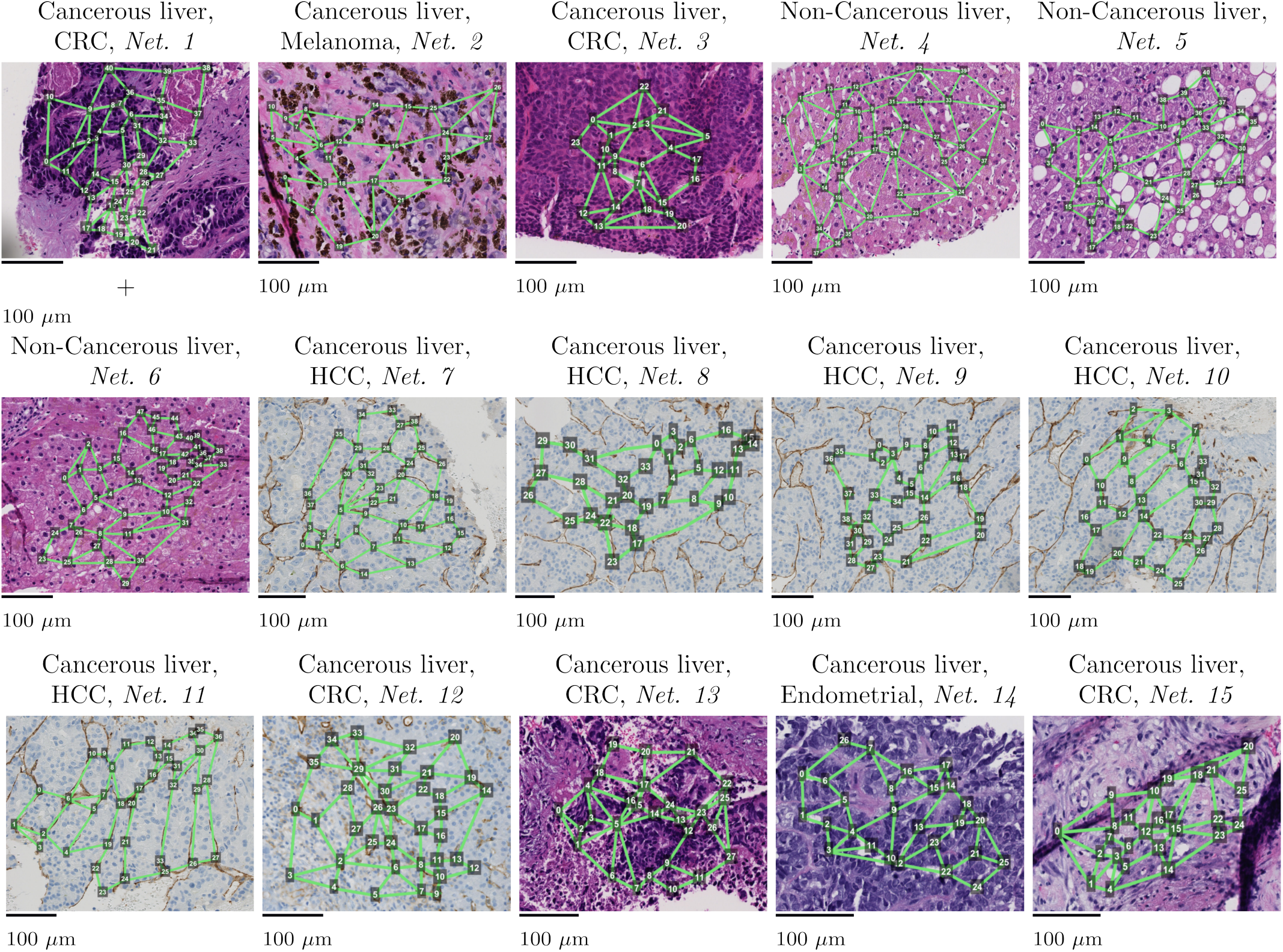
Vascular networks segmented on digitised liver tumor biopsies (resolution: 0.454 *μ*m). Each network is labelled as “Non-Cancerous” or “Cancerous”, depending on whether it was drawn on non-cancerous liver parenchyma or on tumour tissue. For the latter case, the primary cancer is also indicated (CRC stands for Colorectal Cancer, while HCC for Hepatocellular Carcinoma). The non-cancerous networks were drawn on liver tissue found on liver tumour biopsies of patients suffering from Melanoma (n=2) and Ovarian cancer (n=1).

Table 1 reports salient statistics of the vascular networks shown in Fig. 2, related to capillary radii, length, velocity distribution, and number of vascular segments sensed by flowing spins. The table shows that different network morphologies lead to different blood velocity distributions. For example, mean *ν*_*m*_ across VFR realisation can vary from as low as approximately 4 mm/s up to 25 mm/s. This range of variation is mirrored in the average number of capillaries blood travels through during the simulation (*ANB* metric), which varies from just over 12 up to almost 57 segments. Supplementary Fig. 2 shows distributions of *ν*_*m*_, *ν*_*s*_ and *ANB* for all networks, across the 10 different inlet/outlet realisations and given an illustrative input VFR *q*_*in*_ = 3.1*⋅*10^−3^ mm^3^/s. Distributions are skewed, and strong contrasts in terms of *ν*_*m*_, *ν*_*s*_ and *ANB* are seen across networks (e.g., compare Net 3 with Net 4). Supplementary Fig. 3 shows correlation coefficients among all possible pairs of metrics among *ν*_*m*_, *ν*_*s*_, *ANB*, as well as mean segment length *L* and capillary radius *r*. There is a strong, positive correlation between *ν*_*m*_ and *ν*_*s*_ and *ANB* (0.89, 0.82), and a moderate positive correlation between *ν*_*s*_ and *ANB* (0.55). All of *ν*_*m*_, *ν*_*s*_ and *ANB* are negatively correlated with *L* and *r* (strongest correlations between *ANB* and *r*, of -0.93; weakest for *ν*_*s*_ and *L*, of -0.19). Finally, *L* and *r* are positively correlated between each other (correlation of 0.68).

**Table 1:** Summary of vascular networks with corresponding microvascular properties generated for this study. The non-cancerous networks were drawn on non-cancerous liver tissue found on biopsies from melanoma (n=2) and ovarian cancer (n=1) metastases. Mean patient age was 66.2 years. Male = 5, Female = 6. CRC = colorectal cancer, HCC = hepatocellular carcinoma, EC = endometrial cancer.

| Network | Description | $v_m$<br>[mm/s] | $v_s$ [mm/s] | ANB [segments] | $r$ [μm] | $L$ [μm] | Size [mm] |
| --- | --- | --- | --- | --- | --- | --- | --- |
| Net. 1 | Cancerous liver, CRC | 13.76<br>(8.52) | 14.05<br>(9.69) | 46.51 (22.15) | 3.1 (0.9) | 46.24<br>(13.95) | 0.35 |
| Net. 2 | Cancerous liver, melanoma | 11.24<br>(7.01) | 13.69<br>(9.18) | 28.33 (13.92) | 4.02<br>(1.37) | 60.85<br>(21.59) | 0.39 |
| Net. 3 | Cancerous liver, CRC | 24.54<br>(14.9) | 19.76<br>(12.14) | 56.91 (23.3) | 2.45<br>(0.95) | 53.19<br>(21.38) | 0.24 |
| Net. 4 | Non-cancerous liver | 3.92<br>(2.39) | 3.12<br>(1.98) | 12.38 (6.42) | 5.14<br>(1.55) | 90.55<br>(31.33) | 0.50 |
| Net. 5 | Non-cancerous liver | 10.42<br>(6.38) | 8.16<br>(5.01) | 35.95 (17.13) | 3.76<br>(1.03) | 53.03<br>(15.54) | 0.39 |
| Net. 6 | Non-cancerous liver | 10.99<br>(6.79) | 11.05<br>(6.98) | 36.92 (17.55) | 3.43 (0.9) | 48.23<br>(13.26) | 0.42 |
| Net. 7 | Cancerous liver, HCC | 10.54<br>(6.59) | 10.24<br>(6.8) | 33.77 (16.18) | 3.54<br>(1.03) | 60.12<br>(17.81) | 0.36 |
| Net. 8 | Cancerous liver, HCC | 8.42<br>(7.68) | 12.71<br>(10.52) | 13.74 (6.71) | 5.36<br>(1.78) | 70.9<br>(22.43) | 0.60 |
| Net. 9 | Cancerous liver, HCC | 13.92<br>(9.27) | 17.06<br>(12.41) | 32.78 (16.59) | 3.7 (1.05) | 55.28<br>(16.45) | 0.41 |
| Net. 10 | Cancerous liver, HCC | 8.68<br>(5.46) | 9.8 (6.52) | 17.96 (8.71) | 4.71<br>(1.59) | 68.17<br>(21.72) | 0.44 |
| Net. 11 | Cancerous liver, HCC | 19.14<br>(12.03) | 19.92<br>(12.57) | 33.83 (15.22) | 3.37 (1.0) | 61.24<br>(18.89) | 0.44 |
| Net. 12 | Cancerous liver, CRC | 5.49<br>(3.36) | 6.76<br>(4.66) | 19.84 (10.15) | 4.42<br>(1.42) | 60.47<br>(19.98) | 0.38 |
| Net. 13 | Cancerous liver, CRC | 6.44 (4.1) | 6.26<br>(3.98) | 23.84 (11.8) | 4.59<br>(1.66) | 55.5<br>(20.63) | 0.32 |
| Net. 14 | Cancerous liver, EC | 12.45<br>(7.55) | 10.92<br>(6.73) | 48.51 (22.53) | 3.64<br>(1.27) | 45.1<br>(16.13) | 0.29 |
| Net. 15 | Cancerous liver , CRC | 13.54<br>(8.5) | 11.23<br>(6.97) | 44.38 (19.67) | 3.05<br>(1.13) | 50.68<br>(21.07) | 0.32 |

Fig. 3 shows examples of VFR and blood velocity fields reconstructed in two vascular networks with SpinFlowSim, alongside dMRI signals. The two networks were segmented on non-cancerous liver parenchyma of a patient suffering of melanoma (top panel, Net 6) and on metastatic CRC (bottom panel, Net 12). The figure highlights that distributions of VFRs and velocities arise across network segments, owing to their different resistance to flow. The segments with the highest VFRs do not necessarily feature the highest velocities, due to differences in terms of segment diameter. The VFR distributions lead to fast dMRI signal attenuation in both networks, with most of the signal decayed by *b* = 150 s/mm^2^. The signal decay is not mono-exponential (note the log-scale in the y-axis). Oscillatory patterns are also seen as well as some diffusion time dependence, with the dMRI signal decreasing slightly with increasing Δ at fixed *b*.

**Figure 3:**
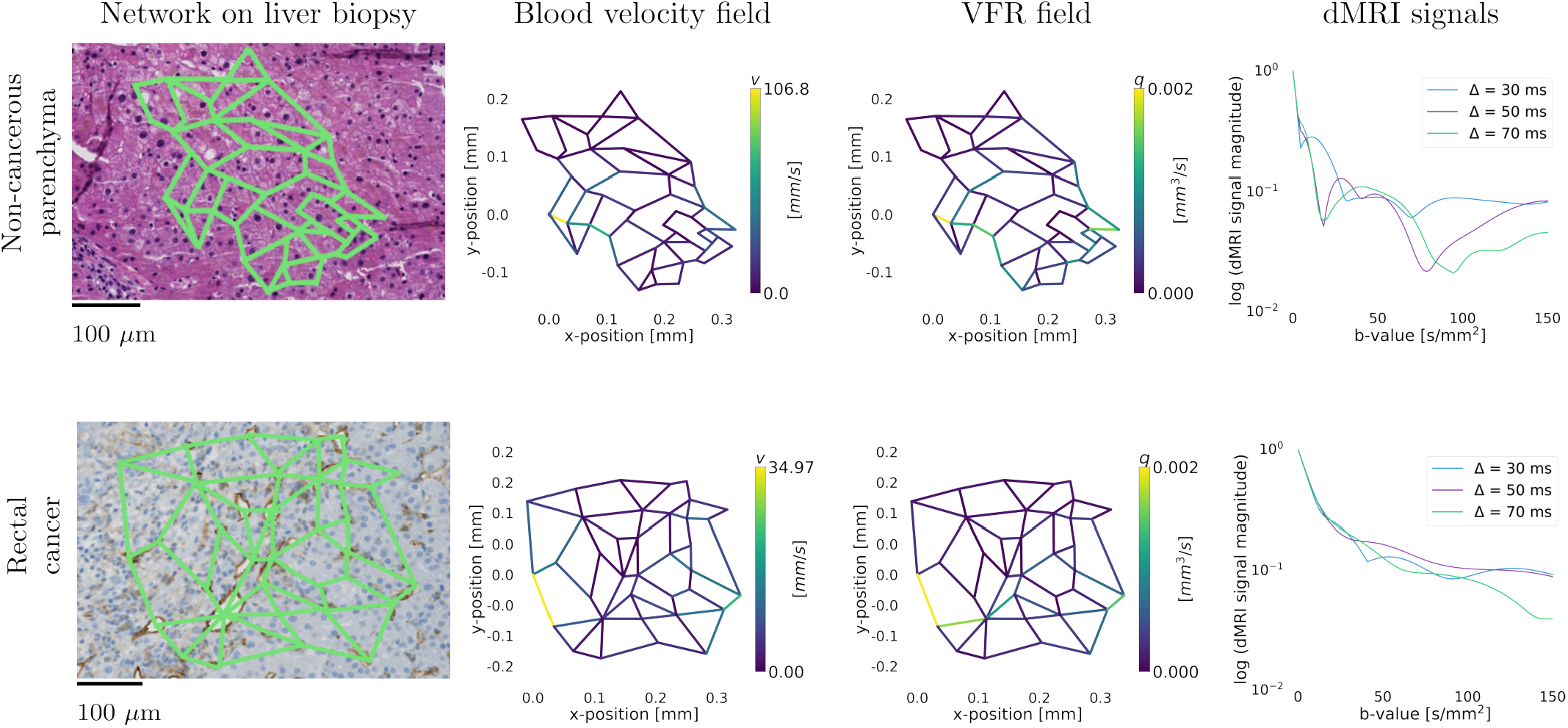
Examples of resolved vascular networks. The top row shows results from a vascular network segmented on a HE-stained non-cancerous liver region, found on a biopsy of a patient with metastatic melanoma (Net 6). The bottom panel shows results from a CD31-stained rectal cancer area (Net 12). From left to right, we show the vascular network, the resolved blood flow velocity field for *q*_*in*_ = 3.1 *⋅* 10^−3^ mm^3^/s, and examples of dMRI signal decay over a range of b-values (0-150 *s*/*mm*^2^) and diffusion times (Δ = {30, 50, 70} ms, *δ* = 20 ms).

Fig. 4 reports on the relationship between *D*^*^ and microvascular properties *ν*_*m*_, *ν*_*s*_ and *ANB. D*^*^ increases with increasing *ν*_*m*_, *ν*_*s*_ and *ANB*. There is a strong positive correlation between *D*^*^ and *ν*_*m*_ and between *D*^*^ and *ANB* (e.g., Spearman’s *r* = 0.85 at Δ = 30 ms for both *ν*_*m*_ and *ANB*), while the correlation is moderate between *D*^*^ and *ν*_*s*_ (*r* = 0.43 at Δ = 30 ms for *ν*_*s*_). The dependence of *D*^*^ on *ν*_*m*_, *ν*_*s*_ and *ANB* is modulated by the diffusion gradient separation Δ, albeit slightly. For all three metrics, *D*^*^ increases with increasing Δ.

**Figure 4:**
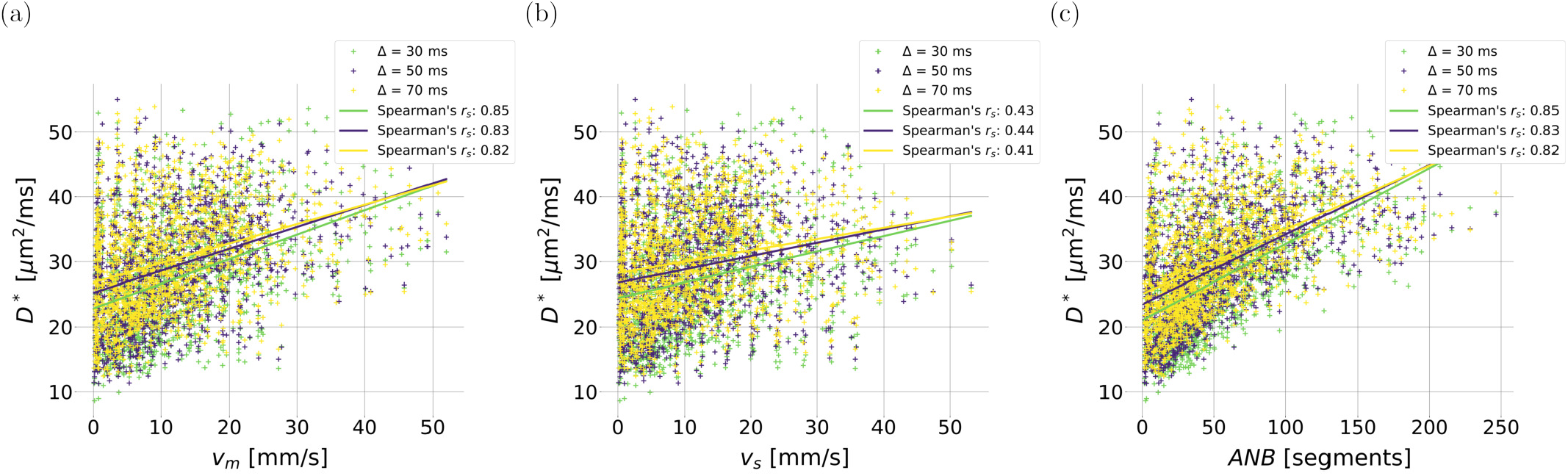
Scatter plots relating *D*^*^ to microvascular properties for different gradient separation Δ and fixed gradient duration *δ* = 20 ms. From left to right: relationship between *D*^*^ and metric *ν*_*m*_ ; *D*^*^ and metric *ν*_*s*_ ; *D*^*^ and metric *ANB*. Spearman’s correlation coefficients between *D*^*^ and each microvascular property are also reported in each plot, for any fixed Δ.

### 3.2 Microvascular property estimation from dMRI

#### 3.2.1 *In silico* estimation

Fig. 5 reports results from *in silico* estimation of *ν*_*m*_, *ν*_*s*_ and *ANB* from noisy vascular signals, synthesised according to protocols “TRSE”, “PGSE” and “richPGSE”. There is a moderate correlation between ground truth and estimated *ν*_*m*_, *ν*_*s*_ and *ANB* values for protocols “PGSE” and “PGSE” (minimum correlation: 0.45 for *ν*_*s*_ for protocol “TRSE”; maximum correlation of 0.65 for *ANB* for protocol “PGSE”). Correlation is instead strong for protocol “richPGSE”, e.g., up to 0.79 for metric *ANB*. As an example, Supplementary Fig. 4 illustrates the complete set of synthetic signals generated for protocol “TRSE” across the 15 segmented networks. The figure highlights that the signal decay spans several order of magnitude: variations in the microvascular characteristics of the networks lead to remarkably different vascular dMRI signals.

**Figure 5:**
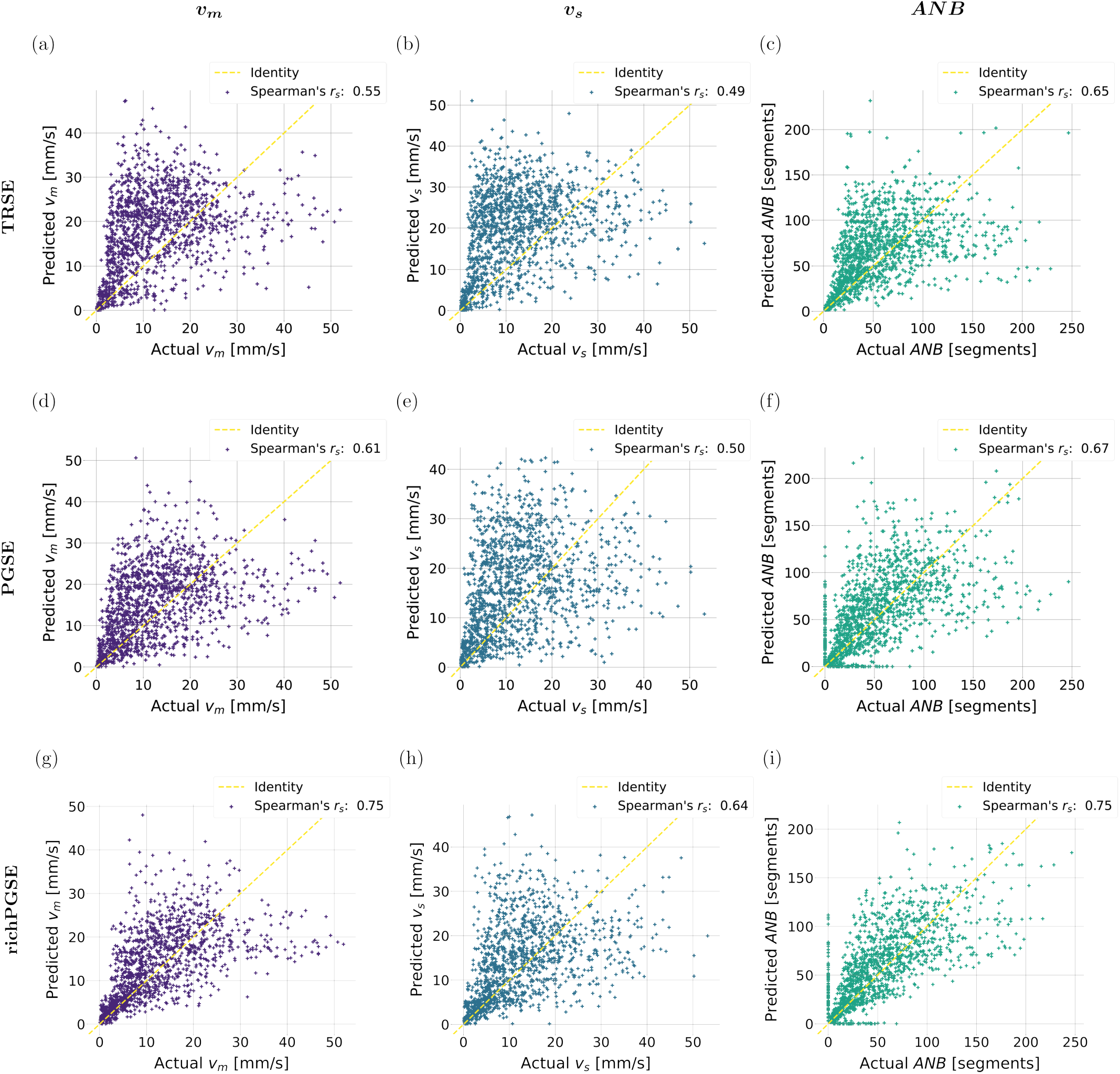
Scatter plots relating estimated and ground truth microvascular parameters in computer simulations. First row (panels (a), (b), (c)): results for protocol “TRSE”. Second row (panels (d), (e), (f)): results for protocol “PGSE”. Third row (panels (g), (h), (i)): results for protocol “richPGSE”. From left to right: results for metric *ν*_*m*_ (panels (a), (d), (g)); results for metric *ν*_*s*_ (panels (b), (e), (h)); results for metric *ANB* (panels (c), (f), (i)). Spearman’s correlation coefficients between estimated and ground truth values are also reported in each plot.

#### 3.2.2 *In vivo* estimation

Fig. 6 shows IVIM metrics *f*_*V*_ and *D*^*^ alongside *ν*_*m*_, *ν*_*s*_ and *ANB* in the liver and spleen of healthy volunteer 1. On visual inspection, *f*_*V*_ and *D*^*^ are systematically higher in the liver than in the spleen. This contrast is mirrored by *ν*_*m*_, *ν*_*s*_ and *ANB*, which are as well higher in the former organ than in the latter. Fig. 7 reports instead mean and standard errors of all metrics within several ROIs (liver, kidney medulla and cortex, and spleen), and in both healthy volunteers. Inter-organ differences are seen, as for example higher *D*^*^, *ν*_*m*_, *ν*_*s*_ and *ANB* in the liver, compared to the spleen. Trends of inter-subject differences are also seen. E.g., in healthy volunteer 1, higher values of all of *f*_*V*_, *D*^*^, *ν*_*m*_, *ν*_*s*_ and *ANB* in the kidney medulla than in the kidney cortex are seen. However, in healthy volunteer 2, *D*^*^ is higher in the cortex than in the medulla, and differences between medulla and cortex among all other metrics are less marked. Intra-/inter-scanner variability is seen in all of *f*_*V*_, *D*^*^, *ν*_*m*_, *ν*_*s*_ and *ANB*.

**Figure 6:**
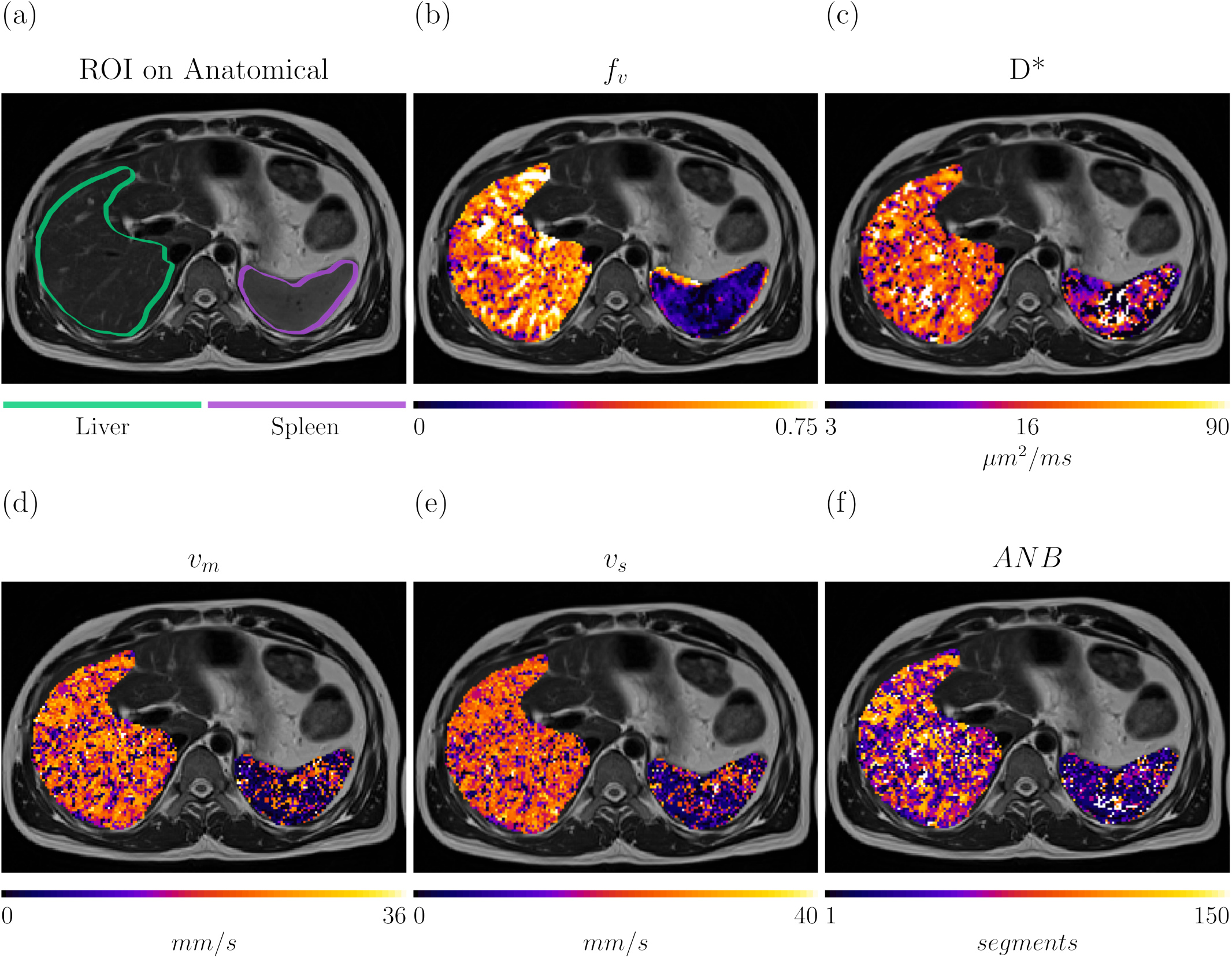
Microvascular maps in a representative healthy volunteer. (a): *b* = 0 image; (b) and (c): IVIM maps *f*_*V*_ and *D*^*^; (d), (e) and (f): microvascular indices *ν*_*m*_, *ν*_*s*_ and *ANB*. In the *b* = 0 image, we highlight the location of the liver and the spleen.

**Figure 7:**
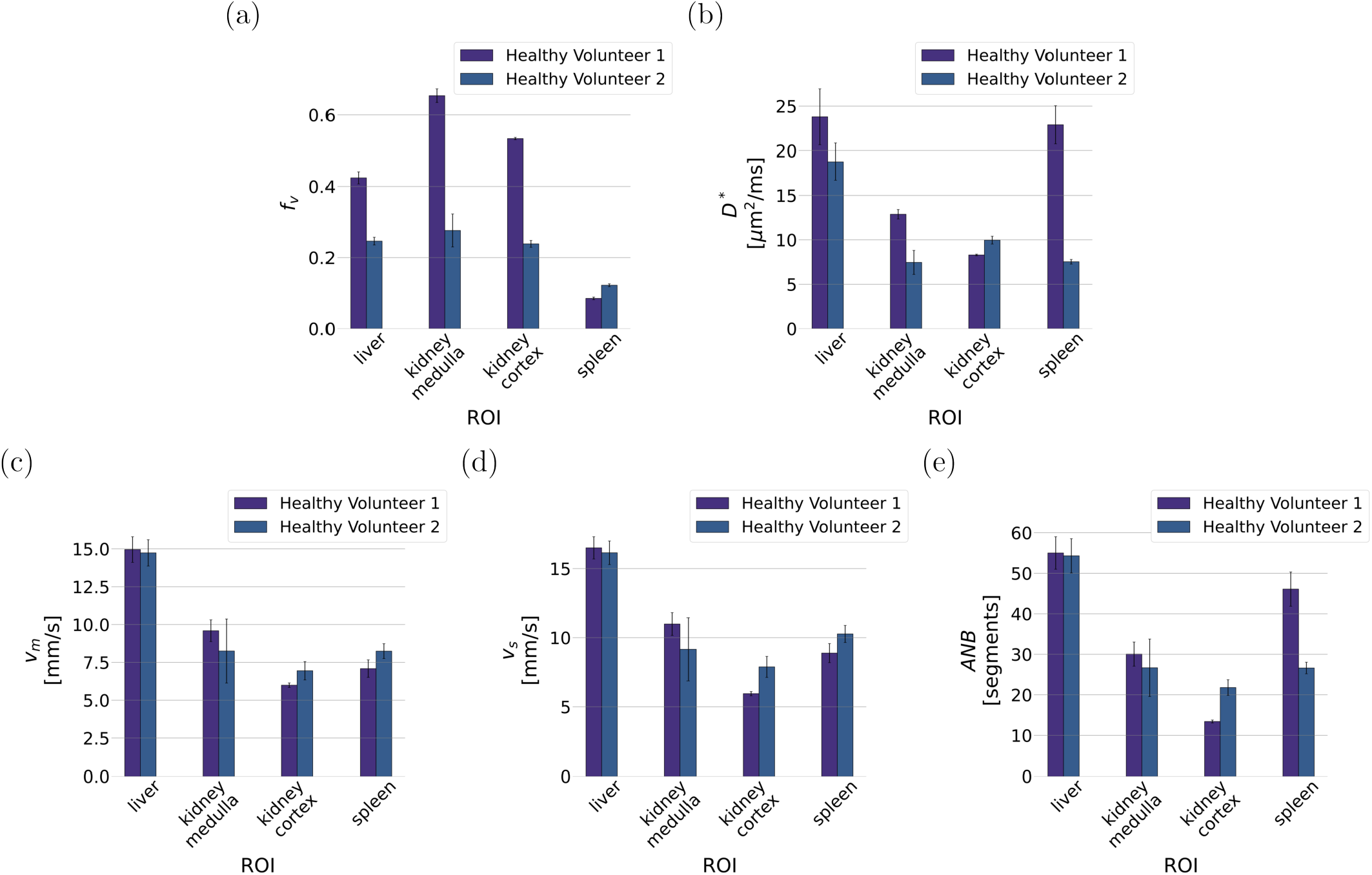
Bar plots reporting mean and standard error of the mean of all microvascular metrics in the different regions-of-interest (ROIs) of both healthy volunteers. (a): trends for metric *f*_*V*_ ; (b): trends for metric *D*^*^; (c): trends for metric *ν*_*m*_; (d): trends for metric *ν*_*s*_; trends for metric *ANB*.

Fig. 8 shows representative microvascular maps in cancer. The figure refers to a colorectal cancer adrenal gland metastasis. Both IVIM metrics *f*_*V*_ and *D*^*^ as well as microvascular *ν*_*m*_, *ν*_*s*_ and *ANB* show intra-tumour contrasts. For example, we observe a core of lower *f*_*V*_ and *D*^*^ as compared to the outer ring of the tumour. This spatial trend is mirrored by metrics *ν*_*m*_, *ν*_*s*_ and *ANB*: the lower *ν*_*m*_, *ν*_*s*_ and *ANB* point towards slower and less variable blood velocity in the core of the tumour, and predict blood to travel through fewer vessel segments, as compared to the outer ring. Overall, *ν*_*m*_, *ν*_*s*_ and *ANB* exhibit similar contrasts among each other, but certain differences are also seen e.g., voxels with high *ν*_*m*_ that do not necessarily show the highest *ANB* values.

**Figure 8.**
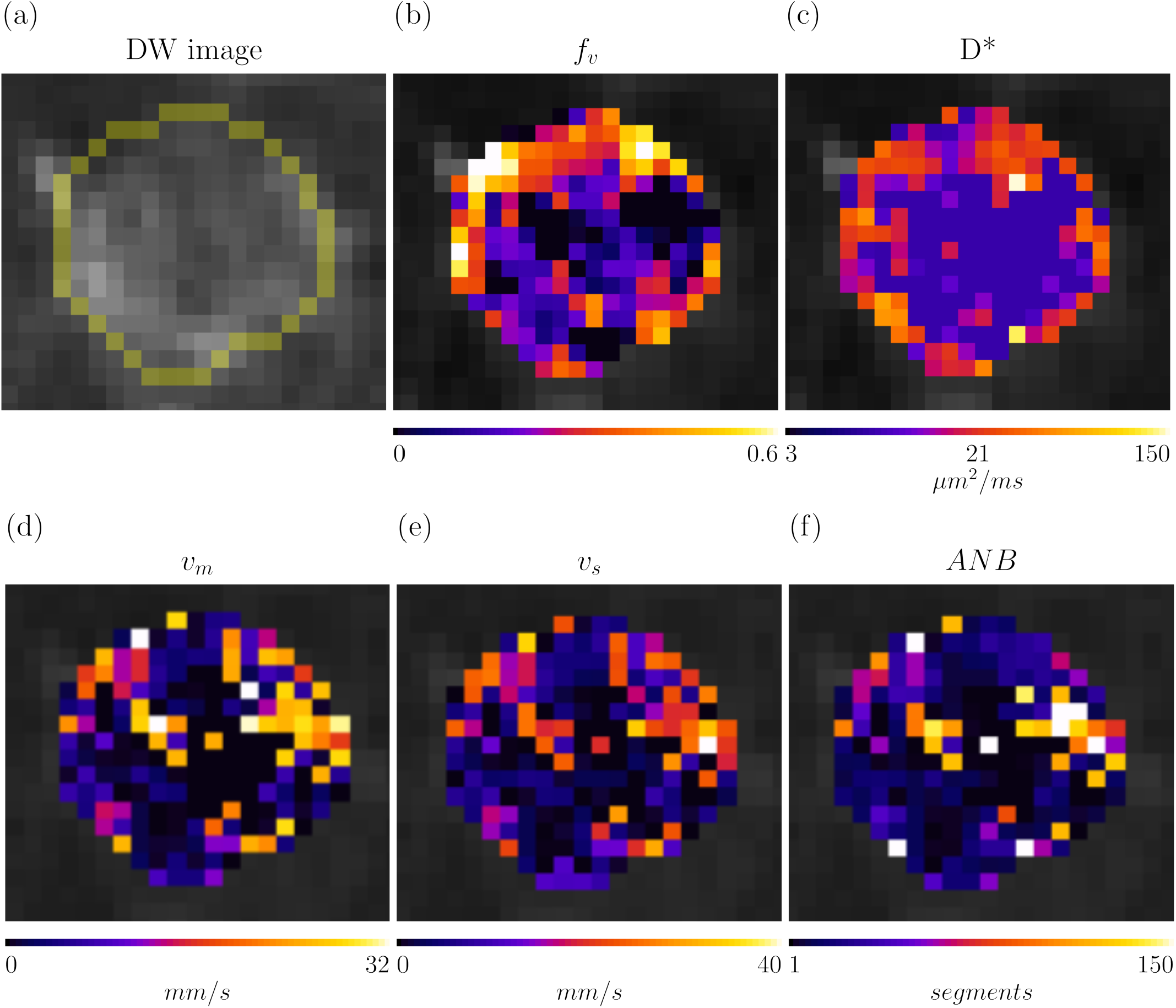
: Parametric maps obtained in an adrenal gland metastasis of a 61 y.o. male patient, suffering from advanced rectal cancer (patient 8, scanned on a 1.5T Siemens Avanto system with a DW TRSE sequence). Top row: *b* = 400 s/mm^2^ image and IVIM maps *f*_*V*_ and *D*^*^. Bottom row: microvascular parameters obtained via simulation-informed model fitting, namely: mean blood velocity *ν*_*s*_, blood velocity standard deviation *ν*_*s*_, and Apparent Network Branching *ANB*.

Table 2 reports mean and standard deviation of all vascular metrics within tumours. The metrics highlight inter-tumour differences in terms of vascularisation, as seen in dMRI. For example, breast cancer metastases feature the highest vascular signal fraction *f*_*V*_ among all tumours. Conversely, the highest *D*^*^ is seen in a lung cancer adrenal gland metastasis (patient 11), which also features the highest *ν*_*m*_, *ν*_*s*_ and *ANB* across the whole cohort. The lowest *D*^*^ is instead seen in liver metastasis of rectal cancer (patient 8), a trend that is mirrored by *ν*_*m*_ and *ANB*, which in this case are the lowest across all tumours.

**Table 2:** Summary of patients’ demographics and key clinical data (primary cancer type, location of the imaged tumours, patients’ sex and approximate age, expressed in 5-years ranges) and trends of microvascular metrics *f*_*V*_, *D*^*^, *ν*_*m*_, *ν*_*s*_ and *ANB* within the manually-segmented tumours (mean and standard deviation). For sex, F indicated female, while M male. Routine IVIM metrics *f*_*V*_ and *D*^*^ were obtained at fixed diffusion time, on the dMRI scan with the shortest TE.

| ID | Primary cancer | Tumour(s) location | Sex | Age | $v_m$<br>[mm/s] | $v_s$<br>[mm/s] | ANB<br>[segments] | $f_V$ | $D^*$<br>[$\mu m^2/ms$] |
| --- | --- | --- | --- | --- | --- | --- | --- | --- | --- |
| Pat. 1 | Endometrial | Uterus | F | 66-70 | 7.34<br>(8.11) | 8.41<br>(8.38) | 28.86<br>(37.71) | 0.11<br>(0.12) | 14.44<br>(22.35) |
| Pat. 2 | Melanoma | Liver | F | 81-85 | 7.12<br>(9.14) | 7.76<br>(8.71) | 31.45<br>(43.12) | 0.20<br>(0.24) | 27.16<br>(43.33) |
| Pat. 3 | Gastric | Soft tissues | M | 61-65 | 7.78<br>(8.90) | 8.53<br>(8.71) | 34.88<br>(46.89) | 0.09<br>(0.09) | 21.09<br>(35.53) |
| Pat. 4 | Melanoma | Liver, lung, pleura | F | 61-65 | 10.72<br>(9.21) | 11.97<br>(9.23) | 42.24<br>(45.49) | 0.19<br>(0.17) | 24.05<br>(33.60) |
| Pat. 5 | Melanoma | Liver | M | 76-80 | 9.96<br>(8.05) | 11.26<br>(7.93) | 39.84<br>(41.39) | 0.04<br>(0.14) | 19.62<br>(21.99) |
| Pat. 6 | Lung | Liver | M | 51-55 | 5.76<br>(8.33) | 6.64<br>(8.26) | 26.88<br>(40.85) | 0.14<br>(0.13) | 15.51<br>(30.47) |
| Pat. 7 | Gastric | Stomach | F | 66-70 | 5.80<br>(7.27) | 7.10<br>(7.52) | 24.76<br>(28.91) | 0.25<br>(0.18) | 13.62<br>(22.17) |
| Pat. 8 | Rectal | Adrenal glands | M | 61-65 | 4.80<br>(7.14) | 5.78<br>(7.58) | 20.80<br>(32.32) | 0.27<br>(0.18) | 10.13<br>(20.32) |
| Pat. 9 | Gastric | Liver | F | 66-70 | 7.49<br>(8.43) | 8.69<br>(8.51) | 31.35<br>(40.54) | 0.22<br>(0.18) | 20.58<br>(35.76) |
| Pat. 10 | Colon | Liver | F | 46-50 | 6.77<br>(8.05) | 7.86<br>(8.21) | 28.21<br>(38.83) | 0.22<br>(0.15) | 17.50<br>(30.04) |
| Pat. 11 | Lung | Adrenal glands | M | 61-65 | 13.37<br>(9.42) | 15.15<br>(9.85) | 55.18<br>(52.69) | 0.26<br>(0.15) | 34.10<br>(44.59) |
| Pat. 12 | Breast | Liver | F | 31-35 | 11.15<br>(10.75) | 11.94<br>(10.48) | 54.71<br>(60.86) | 0.39<br>(0.24) | 14.40<br>(27.20) |
| Pat. 13 | Lung | Adrenal glands | M | 76-80 | 7.77<br>(8.75) | 8.88<br>(8.97) | 32.96<br>(44.13) | 0.15<br>(0.13) | 22.85<br>(36.79) |

Supplementary Fig. 5 reports Spearman’s correlation coefficients between all possible pairs of vascular metrics, as obtained across the 13 cancer patients. IVIM *D*^*^ is significantly, positively correlated with all of *ν*_*m*_, *ν*_*s*_ and *ANB* (*r*_*s*_ = 0.55, *p* = 0.049 with *ν*_*m*_; *r*_*s*_ = 0.57, *p* = 0.044 with *ν*_*s*_; *r*_*s*_ = 0.64, *p* = 0.019 with *ANB*). No significant correlations are instead seen between *f*_*V*_ and any of *ν*_*m*_, *ν*_*s*_ and *ANB. D*^*^ and *f*_*V*_ are weakly, negatively correlated between each other (*r*_*s*_ = -0.25, *p* = 0.394).

## 4 Discussion

### 4.1 Summary and key findings

This work presents SpinFlowSim, an open-source simulator of blood flow based on pipe network analysis. The simulation framework, tailored for the laminar flow regime at the micro-capillary level, enables the synthesis of DW signals for any desired input dMRI gradient waverform. We demonstrate Spin-FlowSim on 15 microvascular networks, reconstructed from biopsies in a variety of liver cancers and in non-cancerous liver parenchyma. These allowed us to simulate micro-perfusion IVIM signals for realistic dMRI protocols, in the low *b* regime. The signals exhibit complex, non-mono-exponential behaviour, pointing towards the co-existence of spin pools experiencing different flow regimes. Key microvascular properties, such as moments of the blood velocity distribution the local network branching sensed by flowing spins, are shown to be encoded in the dMRI signal decay. This fact enabled the design of a strategy for the practical estimation of these properties from any input dMRI signal measurements, given simulations of the corresponding acquisition protocol. We showcase the approach *in silico* and on *in vivo* scans of healthy volunteers and cancer patients, obtaining patterns of microvascular metrics that are plausible with the known anatomy and cancer pathophysiology.

### 4.2 Simulation framework

Our simulator relies on a well-established computational approach for laminar flow characterisation in capillaries. This links the pressure drop across a capillary to the VFR passing through it, via a flow resistance proportionality factor [Schmid et al., 2015, Van et al., 2021]. In this study, as a first proof-of-concept, we borrowed an empirical expression for this resistance from [Blinder et al., 2013], and used the freely available *PySpice* [Salvaire, 2023] package to convert the VFR estimation problem into the analysis of a passive electric circuit. Our strategy, computationally efficient, retrieves the VFR distribution across all segments of a vascular network. These are used to estimate the mean blood velocity in each capillary and, finally, the trajectories of flowing spins, by numerical integration of the velocity field over time. By superimposing arbitrary dMRI gradient wave forms to spins flowing in networks reconstructed from histology, our framework enables the synthesis of realistic IVIM signals, without making assumptions on the specific flow regime in which measurements take place (e.g., diffusive/ballistic [Kennan et al., 1994, Scott et al., 2021]). Our approach offers a practical way to characterise the salient characteristics of micro-capillary perfusion, and its relationship to dMRI. It may therefore play a key role in the development of innovative dMRI methods for vascular characterisation with unprecedented specificity to physiology, urgently needed for non-invasive cancer characterisation.

### 4.3 Vascular networks

We studied HE and CD31-stained histological images from liver tumour biopsies, obtained from cancer patients suffering from advanced solid tumours. From these data, we manually reconstructed 15 2D vascular networks, on which we simulated blood flow by varying the input VFR and the inlet/outlet positions. We characterised the networks in terms of the underlying blood velocity distribution (*ν*_*m*_ and *ν*_*s*_ parameters), and by introducing a metric quantifying the average number of capillary branches spins flow through, referred to as *ANB*. Overall, our simulations generated a total of 1500 network realisations, which provide insight into microvascular blood perfusion. The most important observation is that the networks exhibit blood velocity distributions that can vary considerably from each other, with mean velocity *ν*_*m*_ ranging from approximately 4 to 25 mm/s. This is exemplified, for example, by Net 11 and 12 in Table 1, which feature a mean *ν*_*m*_ of 19 and 5 mm/s, despite exhibiting a similar mean capillary length of circa 60 *μ*m. This finding suggests that, for the typical diffusion times that can be probed in clinical settings (15-65 ms approximately), spins in the vascular compartment likely experience flow regimes that can vary considerably from subject to subject. On the one hand, this implies that hypothesising a specific regime in IVIM modelling (e.g., diffusive versus ballistic [Kennan et al., 1994, Scott et al., 2021]), may not suffice to capture the full complexity of blood micro-perfusion in real-world cohorts. On the other hand, this also implies that remarkably different patterns of vascular dMRI signals are to be expected, even for short, clinically-feasible IVIM dMRI protocols, as exemplified by two examples in Fig. 3. Our simulated signals exhibit complex patterns as a function of the b-value and the diffusion time, e.g., fast decay, typical of the diffusive regime, as well as oscillatory behaviours, as instead expected in the ballistic regime (note that the PGSE signal for a set of uniformly distributed straight capillaries, characterised by the same blood velocity *ν*, is *s* = *sinc*(*γ ν G δ* Δ) [Scott et al., 2021]). Moreover, they also feature a clearly non-mono-exponential behaviour as a function of *b*, pointing again towards the co-existence of different flowing spin pools within the same network, potentially characterised by different flow regimes. All in all, these results suggest that numerical approaches such as SpinFlowSim may lead to more accurate characterisation of unexplored properties ofvascular dMRI signals — e.g., concerning flow anisotropy or apparent pseudo-diffusion and kurtosis tensors, as illustrated in Supplementary Fig. 6 for the apparent pseudo-diffusion tensor in an exemplificative case —, ultimately opening up new opportunities for the development of more specific biomarkers of micro-perfusion.

### 4.4 Microvascular property estimation

We also investigated whether it is possible to use the synthetic signals generated through SpinFlowsim to inform the non-invasive estimation of microvascular properties. For this purpose, we interpolated the full set of paired synthetic signals and microvascular parameters, obtaining numerical forward models that can be fitted through standard NNLS approaches. We specifically investigated the feasibility of estimating *ν*_*m*_, *ν*_*s*_ and *ANB*, since the analysis of simulated signal suggests that even standard PGSE dMRI protocols carry some sensitivity to these properties (note that strong correlations between the vascular ADC *D*^*^ and *ν*_*m*_ and *ANB*, or the moderate correlation between *D*^*^ and *ν*_*s*_, visualised in Fig. 4).

Firstly, we studied *ν*_*m*_, *ν*_*s*_ and *ANB* estimation on noisy *in silico* signals. We considered 3 protocols: two were based on PGSE, with one of these very rich in terms of b-values and diffusion times, and another shorter and clinically feasible. One additional protocol was instead based on DW TRSE, matching that of our *in vivo* data. All protocols point towards the feasibility of estimating *ν*_*m*_, *ν*_*s*_ and *ANB*: we observed strong correlations between ground truth and estimated *ν*_*m*_ and *ANB*, and moderate correlations for *ν*_*s*_. As expected, performances were the highest for the richest protocol, with correlations as high as 0.79 for the *ANB* metric, yet still acceptable for the shorter protocols (e.g., correlation of 0.63 for *ANB* and the TRSE protocol). These promising results, obtained without requiring any explicit analytical modelling of the signal, highlight the potential utility of simulation-informed microvascular property estimation, motivating its testing *in vivo*.

Following *in silico* experiments, we moved on and tested whether *ν*_*m*_, *ν*_*s*_ and *ANB* can also be estimated *in vivo*. For this purpose, we analysed dMRI scans acquired according to the TRSE protocol on two healthy volunteers and in 13 cancer patients. We fitted *ν*_*m*_, *ν*_*s*_ and *ANB* alongside standard IVIM *f*_*V*_ and *D*^*^, and assessed trends qualitatively in several organs in the healthy volunteers, and in the patients’ tumours.

In healthy volunteers, all metrics show high level of variability on visual inspection, which is confirmed by cross-organ trends in Fig. 7. The variability, qualitatively comparable between *f*_*V*_ /*D*^*^ and *ν*_*m*_, *ν*_*s*_ and *ANB*, is in line with the well-known challenge of estimating microvascular property accurately with dMRI [Barbieri et al., 2020]. This finding suggests that more robust parameter estimation procedures may be needed than those used here (e.g., Bayesian fitting or deep learning [Barbieri et al., 2016a, Barbieri et al., 2020]), for the effective deployment of simulation-informed fitting in clinical settings. However, despite the variability, metrics show trends that are compatible with known physiology. For example, in healthy volunteers the liver shows much higher *f*_*V*_, *D*^*^, *ν*_*m*_, *ν*_*s*_ and *ANB* than in the spleen. This finding is plausible considering that the liver is a highly vascularised organ, a blood reservoir receiving approximately 25 % of the cardiac output, despite representing only 2.5 % of the body weight [Lautt, 2010].

We also observe higher *ν*_*m*_, *ν*_*s*_ and *ANB* in the kidney medulla than in the cortex, a finding that may be reflecting their different vascularisation. Regarding kidneys, we do not observe a clear trend in terms of cortex-medulla differences in standard IVIM *f*_*V*_ and *D*^*^ (e.g., *f*_*V*_ is higher in the medulla than in the cortex for both healthy volunteers, while *D*^*^ is in one case higher, and in the other lower). This is in line with recent studies, which have found high variability and strong inter-subject/inter-machine differences of kidney IVIM [Barbieri et al., 2016a,Ljimani et al., 2018, Stabinska et al., 2023].

Finally, we also demonstrated the feasibility of simulation-informed microvascular quantification in a pilot cohort of 13 cancer patients suffering from advanced solid tumours. While this demonstration only represents a first, exploratory proof-of-concept, it serves to highlight that contrasts seen in *ν*_*m*_, *ν*_*s*_ and *ANB* are physiologically plausible, and consistent with patterns seen on *f*_*V*_ and *D*^*^. For example, reduced *ν*_*m*_, *ν*_*s*_ and *ANB* is seen in areas of low *f*_*V*_ and *D*^*^ compatible with reduced perfusion, expected in the tumour core [Karsch-Bluman et al., 2019,Herman et al., 2011], exemplified by Fig. 8. *In vivo, ν*_*m*_, *ν*_*s*_ and *ANB* are positively correlated among each other, and they correlate moderately to strongly to IVIM *D*^*^. These correlations agree with the correlations observed in simulations (compare Supplementary Fig. 3 and 5), and may indicate that *ν*_*m*_, *ν*_*s*_ and *ANB*, while providing complementary information to each other, are sensitive to similar, characteristics of the network morphology. For example, the strong correlation between *ν*_*m*_ and *ν*_*s*_, indicating that higher variability in blood velocity has to be expected as the mean velocity increases, may be a signature of heteroscedasticity of the blood velocity distribution across capillaries.

All in all, our *in vivo* results demonstrate the feasibility of simulation-informed microvascular mapping in dMRI. While further confirmation and more detailed metric characterisation is required in future studies, realistic flow simulations informed by histology may increase the accuracy of dMRI microvascular signal models. Ultimately, these may provide innovative, biologically-specific indices of micro-perfusion, urgently sought for the non-invasive evaluation of cancer neo-angiogenesis, vascular heterogeneity and in treatment during the design of anti-angiogenic drugs.

### 4.5 Methodological considerations and limitations

In this article, we show the potential utility of flow simulations to inform dMRI signal modelling and analysis. We provide a first demonstration, based on a simplified simulation framework as a preliminary proof-of-concept. For example, we rely on an empirical expression for the resistance to flow across a capillary, borrowed from a model of cortical perfusion in the mouse primary sensory cortex [Blinder et al., 2013]. While this model accounts for salient features of blood flow resistance in capillaries (e.g., the effect of an average hematocrit and erythrocyte-wall interactions), a more realistic characterisation of the capillary resistance would be obtained by accounting for the Fåhræus-Lindqvist’s, the Fåhræus’ and the phase separation effects [Schmid et al., 2015, Van et al., 2021]. This would have required the simulation of the propagation of actual erythrocytes through the network, until a steady-state is reached, so that a per-capillary hematocrit (and hence, effective blood viscosity) can be calculated. Here we did not simulate erythrocyte propagation, being this computationally demanding. Nevertheless, we acknowledge that it would enable more realistic representations of flow patterns within micro-capillary networks. We plan to include erythrocyte flow in future work, and also extend SpinFlowSim to account, for example, for oscillatory pressure patterns and vessel deformation, and for fluid exchange between capillaries and the interstitial space.

Furthermore, for this first demonstration, we simulated vascular dMRI signals on 2D capillary networks. While SpinFlowSim is designed to work with generic 3D networks, here we focussed on 2D representations due to the availability of 2D data (i.e., HE and CD31-stained biopsies). We accounted for this by averaging synthetic dMRI signals generated for two, orthogonal, in-plane gradient directions. However, in future we plan to increase the fidelity of our flow simulations by reconstructing 3D networks.

Related to the point above, the vascular networks reconstructed from histology for this article were obtained at the capillary level. Therefore, our synthetic signals may not be representative of larger vessels, including smaller feeding arterioles and small veins or venules. This implies that maps of *ν*_*m*_, *ν*_*s*_ and *ANB* from our approach has to be taken with care in presence of larger vessels. In future, we plan to expand our vascular signal dictionary to include realisations of larger vessels, and thus improve the generalisability of our simulation-informed fitting.

Another point to acknowledge is that in this study we focussed on the characterisation of vascular dMRI signals, and devised a simulation-informed fitting procedure requiring pure vascular signals as input. For this reason, the analysis of *in vivo* signals required disentangling vascular from extra-vascular tissue signals, since low *b* measurements include contributions from both. This was achieved by extrapolating and ADC fit performed on b-values with negligible vascular signal contribution, and thus required identifying a b-value threshold. An approach of this type, i.e., splitting the vascular-tissue signal characterisation in two steps, is sometimes referred to as *segmented IVIM fitting* [Gurney-Champion et al., 2018,Wang et al., 2021]. While segmented fitting reduces the variability of vascular metrics, since it avoids the challenging, joint estimation of vascular and tissue properties [Barbieri et al., 2020], it may lead to biases in *f*_*V*_ estimates, since *f*_*V*_ may depend, at least slightly, on the b-value threshold. In future, we plan to improve the simulation-informed fitting performed here, by employing more advanced estimation techniques for the joint computation of vascular and tissue properties.

Lastly, we acknowledge that the results reported here should be confirmed by future studies. These would require the acquisition of data from additional healthy volunteers and from larger patient cohorts, and should include diffusion images from different MRI scanners and from more advanced dMRI protocols. Here, we used a simple acquisition scheme in the low *b* regime (*b* = 0 and *b* = {50, 100} s/mm^2^ at 3 diffusion times). However, the accurate characterisation of the complex signal patterns arising from microvasculature would likely require denser *b* samplings. Similarly, higher image quality and increased sensitivity to micro-perfusion could also be achieved, for example, by improving the robustness of the dMRI acquisition with cardiac/respiratory gating, or by employing flow-compensated [Wetscherek et al., 2015] gradient wave forms, or advanced b-tensor encoding [Nilsson et al., 2021].

## 5 Conclusions

SpinFlowSim, our open-source, freely-available python simulator of blood micro-perfusion in capillaries, enables the synthesis and characterisation of realistic microvascular dMRI signals. Perfusion simulations in vascular networks reconstructed from histology may inform the non-invasive, numerical estimation of innovative microvascular properties through dMRI, whose feasibility is demonstrated herein *in vivo* in healthy subjects and in cancer patients.

## Data Availability

SpinFlowSim is made freely available as a GitHub repository at the permanent address:
https://github.com/radiomicsgroup/SpinFlowSim. The repository includes the 15 vascular networks presented in this study that can be used to generate synthetic signals to inform model fitting. The code for simulation-informed fitting is freely available as part of BodyMRITools at the permanent address: https://github.com/fragrussu/bodymritools (script mri2micro dictml.py). The in vivo human data cannot be made freely available at this stage due to ethical restrictions.

## Acknowledgements

VHIO would like to acknowledge: the State Agency for Research (Agencia Estatal de Investigación) for the financial support as a Center of Excellence Severo Ochoa (CEX2020-001024-S/AEI/10.13039/501100011033), the Cellex Foundation for providing research facilities and equipment and the CERCA Programme from the Generalitat de Catalunya for their support on this research. This research has been supported by PREdICT, sponsored by AstraZeneca. This study has been cofunded by the European Regional Development Fund/European Social Fund ‘A way to make Europe’ (to R.P.L.), and by the Comprehensive Program of Cancer Immunotherapy and Immunology (CAIMI), funded by the Banco Bilbao Vizcaya Argentaria Foundation Foundation (FBBVA, grant 89/2017). R.P.L is supported by the “la Caixa” Foundation CaixaResearch Advanced Oncology Research Program, the Prostate Cancer Foundation (18YOUN19), a CRIS Foundation Talent Award (TALENT19-05), the FERO Foundation through the XVIII Fero Fellowship for Oncological Research, the Instituto de Salud Carlos III-Investigación en Salud (PI18/01395 and PI21/01019), the Asociación Española Contra el Cancer (AECC) (PRYCO211023SERR) and the Generalitat de Catalunya Agency for Management of University and Research Grants of Catalonia (AGAUR) (2023PROD00178). The project that gave rise to these results received the support of a fellowship from “la Caixa” Foundation (ID 100010434). The fellowship code is “LCF/BQ/PR22/11920010” (funding F.G, A.V., and A.G.). This research has received support from the Beatriu de Pinós Postdoctoral Program from the Secretariat of Universities and Research of the Department of Business and Knowledge of the Government of Catalonia, and the support from the Marie Sklodowska-Curie COFUND program (BP3, contract number 801370; reference 2019 BP 00182) of the H2020 program (to K.B.). M.P. is supported by the UKRI Future Leaders Fellowship MR/T020296/2. A.G. is supported by a Severo Ochoa PhD fellowship (PRE2022-102586). The authors are thankful to the Vall d’Hebron Radiology department and to the ASCIRES CETIR clinical team for their assistance with MRI acquisitions, and to the GE/Siemens clinical scientists for their support with diffusion sequence characterisation.

## Data and code availability

*SpinFlowSim* is made freely available as a GitHub repository at the permanent address: https://github.com/radiomicsgroup/SpinFlowSim. The repository includes the 15 vascular networks presented in this study that can be used to generate synthetic signals to inform model fitting. The code for simulation-informed fitting is freely available as part of *BodyMRITools* at the permanent address: https://github.com/fragrussu/bodymritools (script *mri2micro_dictml*.*py*). The *in vivo* human data cannot be made freely available at this stage due to ethical restrictions.

## Credit author statement

Conceptualization: AV, FG, RPL, DSN, EF, MP. Data curation: AV, FG, RPL, KB, AG, GS, PN, RT, EG. Formal analysis: AV, FG. Funding acquisition: RPL, EG, PN, RT, FG. Investigation: AV, FG, KB, GS, SS, MV. Methodology: AV, FG, AG, RPL, MP, DSN, EF. Project administration: AV, FG, KB, RPL, EG, PN, RT. Resources: AV, FG, RPL, AG, EG, PN, RT, ME, NR. Software: AV, FG, AG. Supervision: FG, RPL, RSL, DSN, EF, MP. Validation: AV, FG. Visualization: AV. Writing – original draft: AV, FG. Writing – review and editing: all authors.

## Supplementary Material

**Supplementary Fig. 1:**
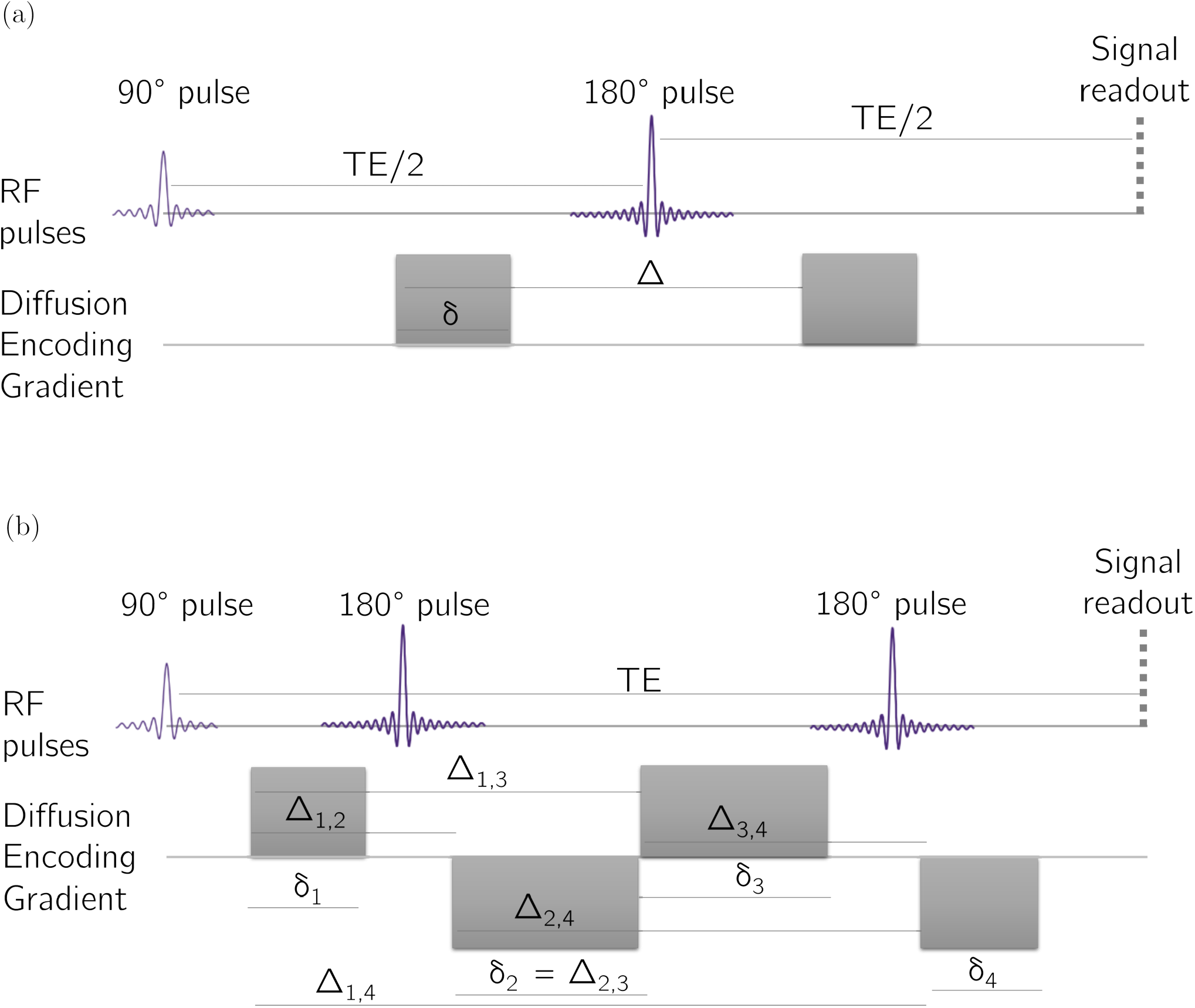
Illustration of the PGSE (panel (a)) and DW-TRSE sequences used in simulations and implemented for actual *in vivo* data acquisition.

**Supplementary Fig. 2:**
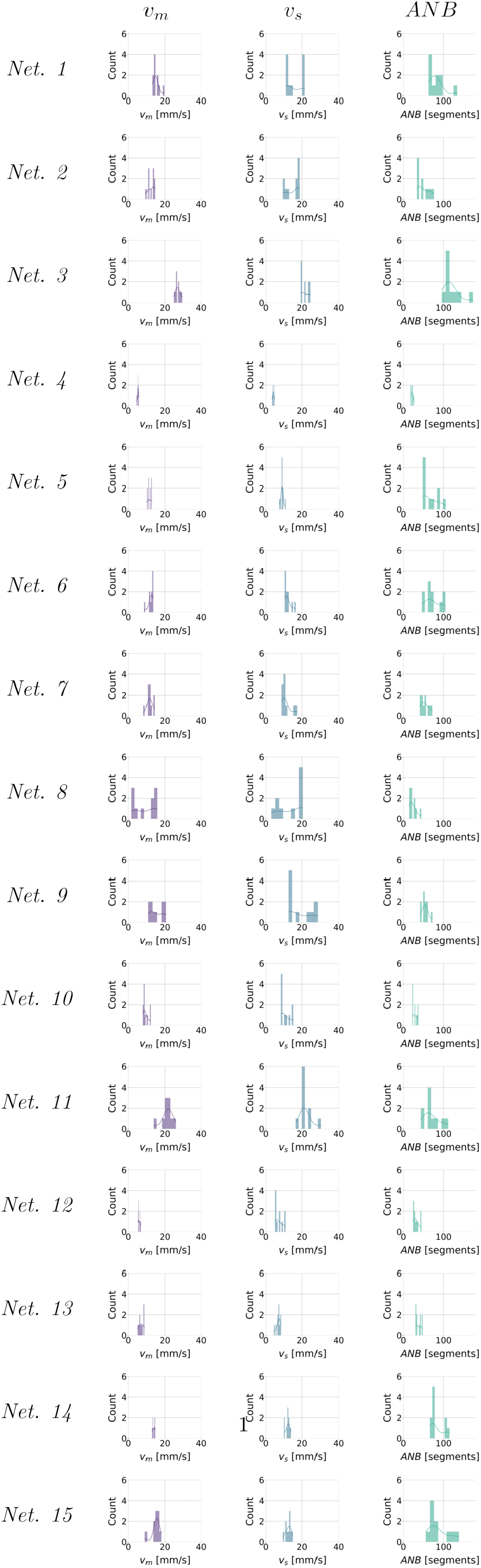
Histograms describing *ν*_*m*_, *ν*_*s*_ and *ANB* distributions obtained across the 10 realisations of each vascular network for an input volumetric flow rate of *q*_*in*_ = 3.1 *⋅* 10^−3^ mm^3^/s.

**Supplementary Fig. 3:**
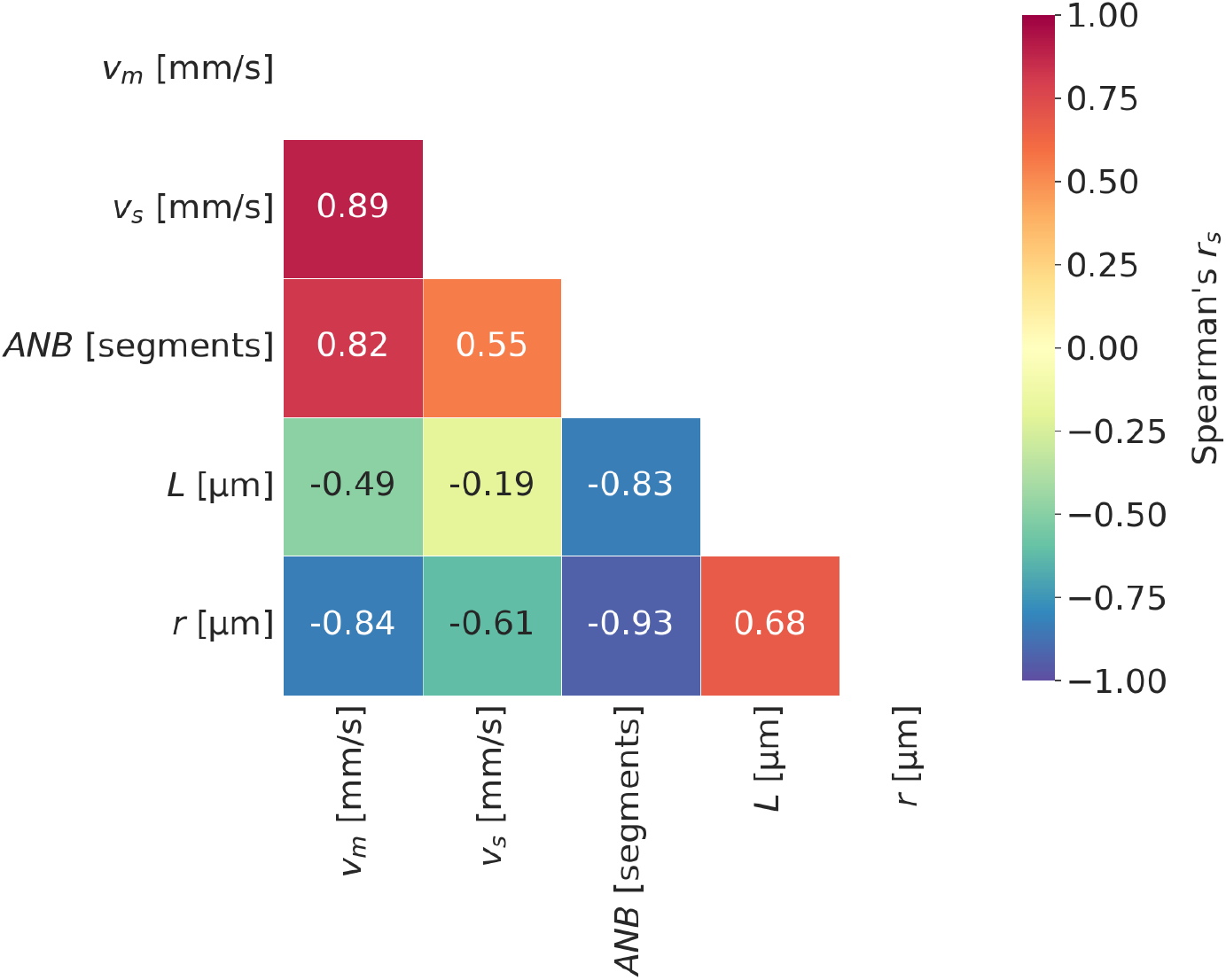
Spearman’s correlation coefficients, in form of a correlation matrix, for parameters *ν*_*m*_ (mean blood velocity), *ν*_*s*_ (standard deviation of blood velocities), *ANB* (apparent network branching), *L* (mean segment length) and *r* (mean radius length) from the 15 vascular networks segmented for this study. Values of *ν*_*m*_, *ν*_*s*_, *ANB, L* and *r* used to compute the matrix are reported in Table 1 (mean values).

**Supplementary Fig. 4:**
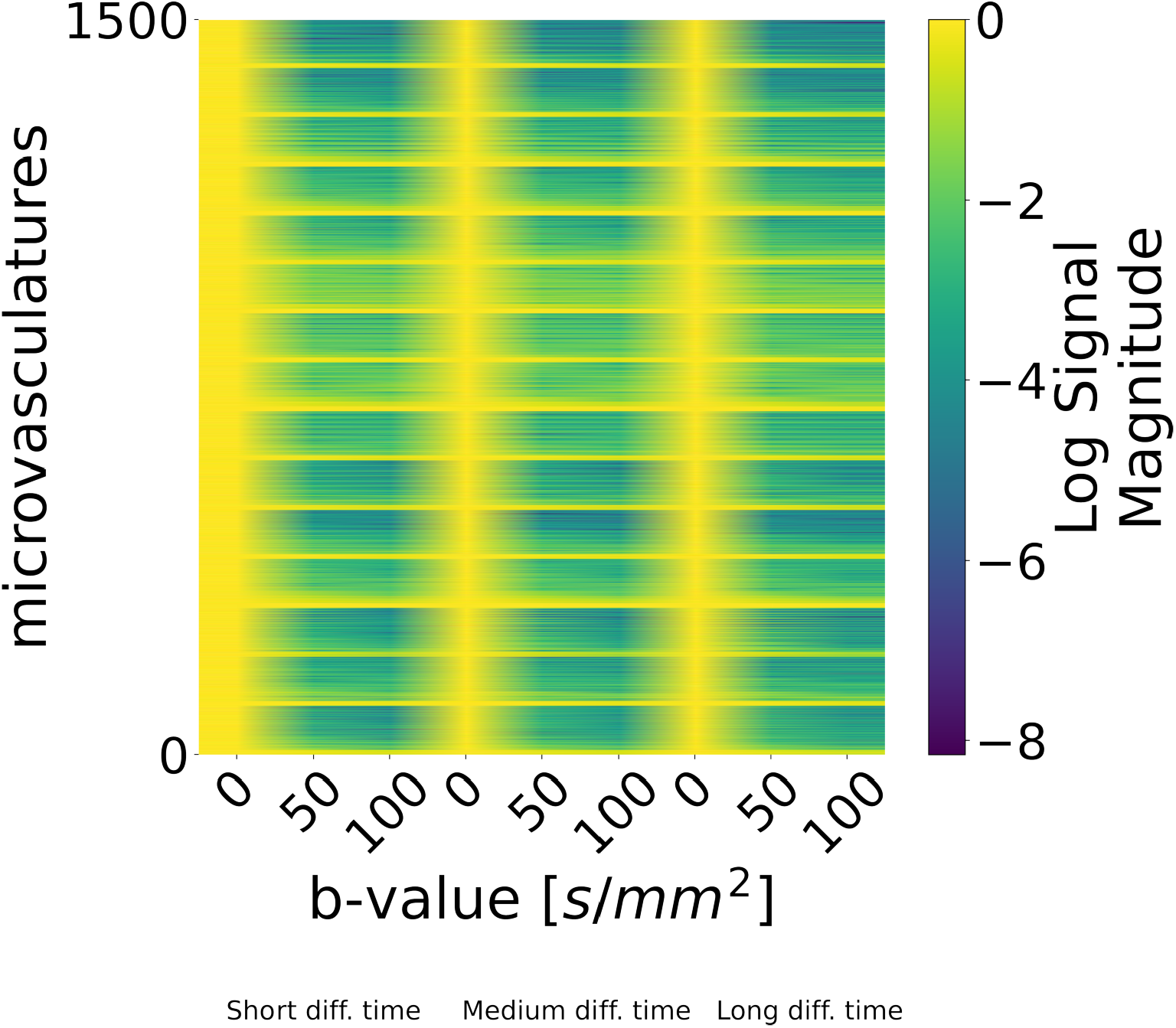
Synthetic signals generated according to *in silico* protocol “TRSE”, and used for learning numerical forward signal models.

**Supplementary Fig. 5:**
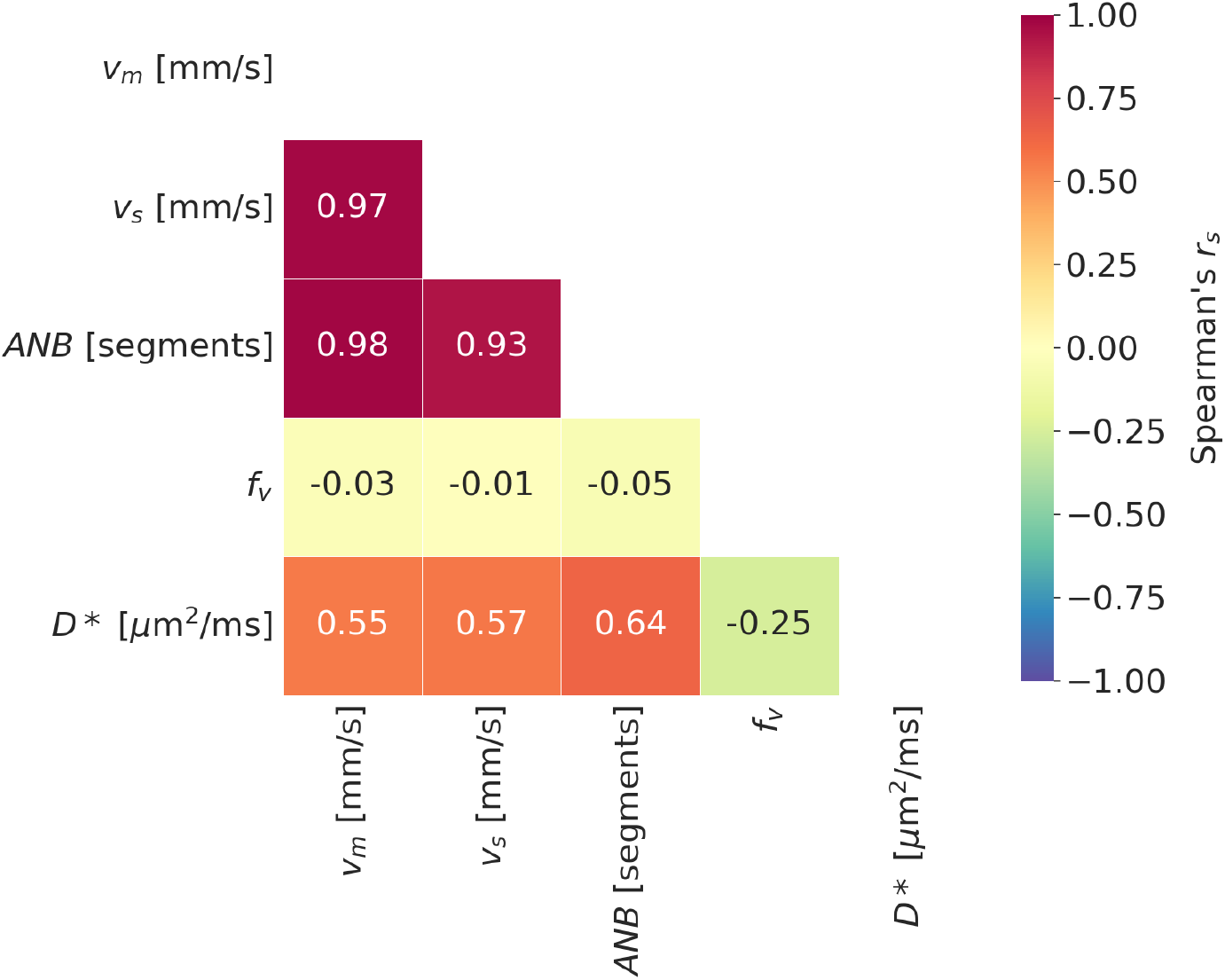
Spearman’s correlation coefficients, in form of a correlation matrix, for parameters *ν*_*m*_ (mean blood velocity), *ν*_*s*_ (standard deviation of blood velocities), *ANB* (apparent network branching), *f*_*V*_ (IVIM vascular signal fraction) and *D*^*^ (vascular ADC) from the 13 cancer patients included in this study. Values of *ν*_*m*_, *ν*_*s*_, *ANB, f*_*V*_ and *D*^*^ used to compute the matrix are reported in Table 2 (mean values).

**Supplementary Fig. 6:**
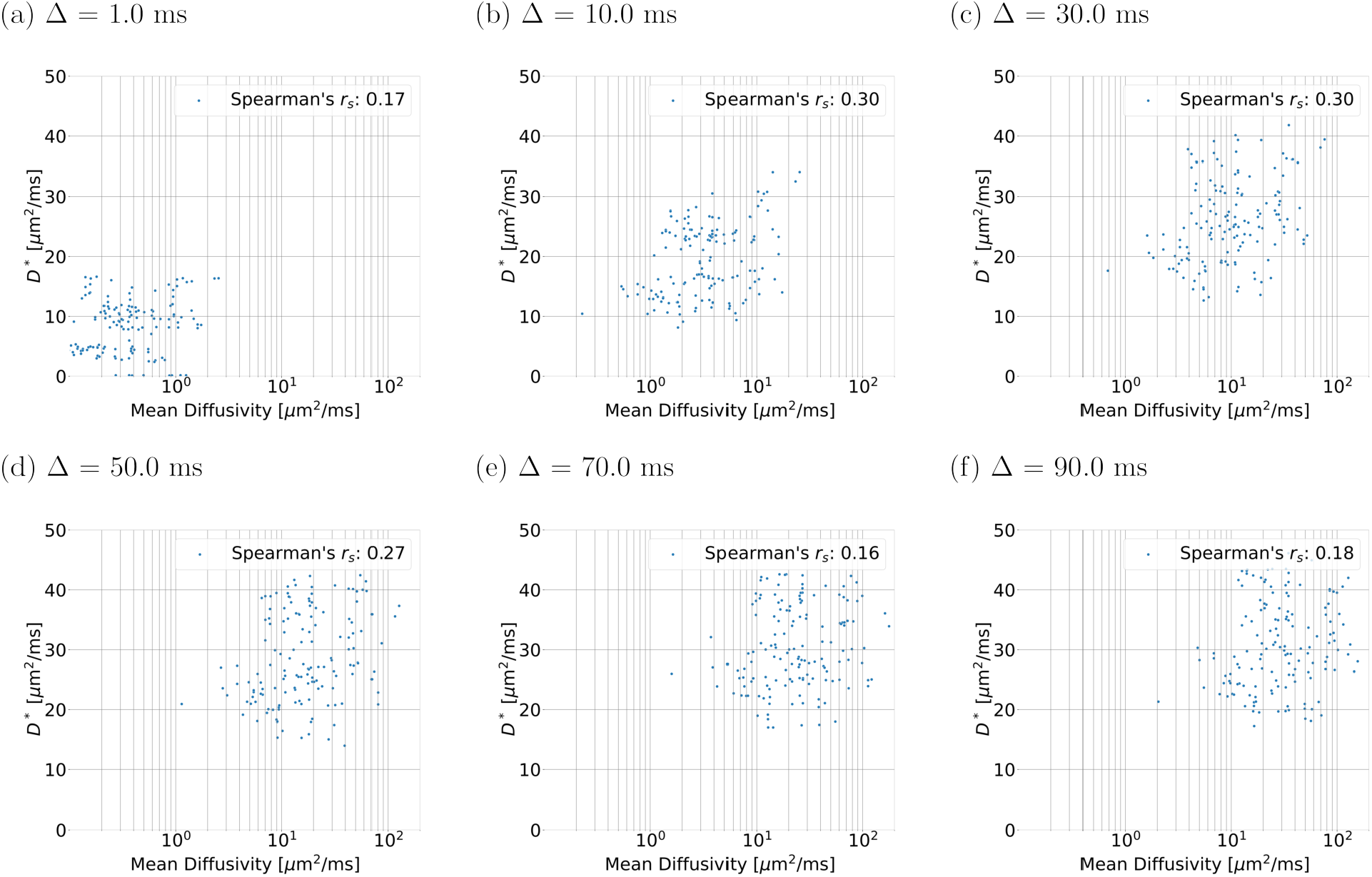
Scatter plots illustrating the relationship between routine IVIM D^*^ and the mean diffusivity of the apparent diffusion tensor 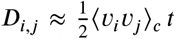 for spins flowing in the ballistic regime [Kennan et al., 1994]. Above, *t* is the diffusion time (*t* ≈ Δ for PGSE in the narrow-pulse regime), while *i* = 1, 2, 3 and *j* = 1, 2, 3 iterate over the *x, y* and *z* components of the capillary velocity vectors, and ⟨*⋅* ⟩c; is the ensemble average over the capillary set. The figure was obtained by pooling together data points for all 100 realisations of the 15 vascular networks drawn for this study, with *q*_*in*_ = 3.1 *⋅* 10^−3^ mm^3^/s and *δ* = 0.1 ms. From top to bottom, clock-wise: (a): Δ = 1 ms; (b): Δ = 10 ms; (c): Δ = 30 ms; (d): Δ = 50 ms; (e): Δ = 70 ms; (f): Δ = 90 ms. The mean diffusivity of *D*_*i,j*_, by construction, increases with increasing diffusion time, a fact that is replicated by the D^*^ metric obtained directly from our synthetic signals. However, there is a relatively weak-to-moderate correlation between D^*^ and the apparent diffusion tensor mean diffusivity, suggesting that complex features of the vascular signal (which influence D^*^ estimation) may not be captured by simple analytical expressions for diffusion cumulants derived in specific flow regimes.

## References

Andersson, J. L., Skare, S., and Ashburner, J. (2003). How to correct susceptibility distortions in spin-echo echo-planar images: application to diffusion tensor imaging. Neuroimage, 20(2):870–888.

Barbieri, S., Donati, O. F., Froehlich, J. M., and Thoeny, H. C. (2016a). Comparison of intravoxel incoherent motion parameters across MR imagers and field strengths: evaluation in upper abdominal organs. Radiology, 279(3):784–794. Publisher: Radiological Society of North America.

Barbieri, S., Donati, O. F., Froehlich, J. M., and Thoeny, H. C. (2016b). Impact of the calculation algorithm on biexponential fitting of diffusion-weighted MRI in upper abdominal organs. Magnetic resonance in medicine, 75(5):2175–2184. Publisher: Wiley Online Library.

Barbieri, S., Gurney-Champion, O. J., Klaassen, R., and Thoeny, H. C. (2020). Deep learning how to fit an intravoxel incoherent motion model to diffusion-weighted MRI. Magnetic Resonance in Medicine, 83(1):312–321.

Blinder, P., Tsai, P. S., Kaufhold, J. P., Knutsen, P. M., Suhl, H., and Kleinfeld, D. (2013). The cortical angiome: an interconnected vascular network with noncolumnar patterns of blood flow. Nature Neuroscience, 16(7):889–897.

Buizza, G., Paganelli, C., Ballati, F., Sacco, S., Preda, L., Iannalfi, A., Alexander, D. C., Baroni, G., and Palombo, M. (2021). Improving the characterization of meningioma microstructure in proton therapy from conventional apparent diffusion coefficient measurements using monte carlo simulations of diffusion MRI. Medical Physics, 48(3):1250–1261. Publisher: Wiley Online Library.

Dappa, E., Elger, T., Hasenburg, A., Düber, C., Battista, M. J., and Hötker, A. M. (2017). The value of advanced MRI techniques in the assessment of cervical cancer: a review. Insights into imaging, 8:471–481. Publisher: Springer.

Fieremans, E. and Lee, H.-H. (2018). Physical and numerical phantoms for the validation of brain microstructural MRI: A cookbook. NeuroImage, 182:39–61.

Fokkinga, E., Hernandez-Tamames, J. A., Ianus, A., Nilsson, M., Tax, C. M., Perez-Lopez, R., and Grussu, F. (2023). Advanced diffusion-weighted MRI for cancer microstructure assessment in body imaging, and its relationship with histology. Journal of Magnetic Resonance Imaging.

Ginsburger, K., Matuschke, F., Poupon, F., Mangin, J.-F., Axer, M., and Poupon, C. (2019). MEDUSA: A GPU-based tool to create realistic phantoms of the brain microstructure using tiny spheres. NeuroImage, 193:10–24. Publisher: Elsevier.

Gurney-Champion, O. J., Klaassen, R., Froeling, M., Barbieri, S., Stoker, J., Engelbrecht, M. R., Wilmink, J. W., Besselink, M. G., Bel, A., Van Laarhoven, H. W., et al. (2018). Comparison of six fit algorithms for the intra-voxel incoherent motion model of diffusion-weighted magnetic resonance imaging data of pancreatic cancer patients. PloS one, 13(4):e0194590.

Hall, M. G. and Alexander, D. C. (2009). Convergence and parameter choice for monte-carlo simulations of diffusion MRI. IEEE transactions on medical imaging, 28(9):1354–1364. Publisher: IEEE.

Herman, A. B., Savage, V. M., and West, G. B. (2011). A quantitative theory of solid tumor growth, metabolic rate and vascularization. PloS one, 6(9):e22973.

Huinen, Z. R., Huijbers, E. J., van Beijnum, J. R., Nowak-Sliwinska, P., and Griffioen, A. W. (2021). Anti-angiogenic agents—overcoming tumour endothelial cell anergy and improving immunotherapy outcomes. Nature Reviews Clinical Oncology, 18(8):527–540. Publisher: Nature Publishing Group UK London.

Iima, M., Kataoka, M., Kanao, S., Onishi, N., Kawai, M., Ohashi, A., Sakaguchi, R., Toi, M., and Togashi, K. (2018). Intravoxel incoherent motion and quantitative non-gaussian diffusion MR imaging: evaluation of the diagnostic and prognostic value of several markers of malignant and benign breast lesions. Radiology, 287(2):432–441. Publisher: Radiological Society of North America.

Ivanov, K., Kalinina, M., and Levkovich, Y. I. (1981). Blood flow velocity in capillaries of brain and muscles and its physiological significance. Microvascular research, 22(2):143–155.

Jayson, G. C., Kerbel, R., Ellis, L. M., and Harris, A. L. (2016). Antiangiogenic therapy in oncology: current status and future directions. The Lancet, 388(10043):518–529. Publisher: Elsevier.

Karsch-Bluman, A., Feiglin, A., Arbib, E., Stern, T., Shoval, H., Schwob, O., Berger, M., and Benny, O. (2019). Tissue necrosis and its role in cancer progression. Oncogene, 38(11):1920–1935.

Kellner, E., Dhital, B., Kiselev, V. G., and Reisert, M. (2016). Gibbs-ringing artifact removal based on local subvoxel-shifts. Magnetic Resonance in Medicine, 76(5):1574–1581.

Kennan, R. P., Gao, J.-H., Zhong, J., and Gore, J. C. (1994). A general model of microcirculatory blood flow effects in gradient sensitized mri. Medical physics, 21(4):539–545.

Kiselev, V. G. (2017). Fundamentals of diffusion MRI physics. NMR in Biomedicine, 30(3):e3602.

Késmárky, G., Kenyeres, P., Rábai, M., and Tóth, K. (2008). Plasma viscosity: a forgotten variable. Clinical hemorheology and microcirculation, 39(1):243–246. Publisher: IOS Press.

Lautt, W. W. (2010). Hepatic circulation: physiology and pathophysiology.

Le Bihan, D. (2019). What can we see with IVIM MRI? Neuroimage, 187:56–67. Publisher: Elsevier.

Le Bihan, D., Breton, E., Lallemand, D., Grenier, P., Cabanis, E., and Laval-Jeantet, M. (1986). MR imaging of intravoxel incoherent motions: application to diffusion and perfusion in neurologic disorders. Radiology, 161(2):401–407.

Le Bihan, D. and Turner, R. (1992). The capillary network: a link between IVIM and classical perfusion. Magnetic Resonance in Medicine, 27(1):171–178.

Lee, H.-H., Fieremans, E., and Novikov, D. S. (2021). Realistic microstructure simulator (RMS): Monte carlo simulations of diffusion in three-dimensional cell segmentations of microscopy images. Journal of neuroscience methods, 350:109018. Publisher: Elsevier.

Ljimani, A., Lanzman, R. S., Müller-Lutz, A., Antoch, G., and Wittsack, H.-J. (2018). Nongaussian diffusion evaluation of the human kidney by padé exponent model. Journal of Magnetic Resonance Imaging, 47(1):160–167.

Morelli, L., Palombo, M., Buizza, G., Riva, G., Pella, A., Fontana, G., Imparato, S., Iannalfi, A., Orlandi, E., Paganelli, C., and others (2023). Microstructural parameters from DW-MRI for tumour characterization and local recurrence prediction in particle therapy of skullbase chordoma. Medical Physics. Publisher: Wiley Online Library.

Munn, L. L. (2003). Aberrant vascular architecture in tumors and its importance in drug-based therapies. Drug discovery today, 8(9):396–403. Publisher: Elsevier.

Nedjati-Gilani, G. L., Schneider, T., Hall, M. G., Cawley, N., Hill, I., Ciccarelli, O., Drobnjak, I., Wheeler-Kingshott, C. A. G., and Alexander, D. C. (2017). Machine learning based compartment models with permeability for white matter microstructure imaging. NeuroImage, 150:119–135. Publisher: Elsevier.

Nguyen, H., Li, J., Grebenkov, D., Le Bihan, D., and Poupon, C. (2014). Parameters estimation from the diffusion mri signal using a macroscopic model. In Journal of Physics: Conference Series, volume 490, page 012117. IOP Publishing.

Nilsson, M., Alerstam, E., Wirestam, R., Sta, F., Brockstedt, S., Lätt, J., and others (2010). Evaluating the accuracy and precision of a two-compartment kärger model using monte carlo simulations. Journal of Magnetic Resonance, 206(1):59–67. Publisher: Elsevier.

Nilsson, M., Eklund, G., Szczepankiewicz, F., Skorpil, M., Bryskhe, K., Westin, C.-F., Lindh, C., Blomqvist, L., and Jäderling, F. (2021). Mapping prostatic microscopic anisotropy using linear and spherical b-tensor encoding: a preliminary study. Magnetic resonance in medicine, 86(4):2025–2033.

Palombo, M., Alexander, D. C., and Zhang, H. (2019). A generative model of realistic brain cells with application to numerical simulation of the diffusion-weighted MR signal. NeuroImage, 188:391–402. Publisher: Elsevier.

Panagiotaki, E., Schneider, T., Siow, B., Hall, M. G., Lythgoe, M. F., and Alexander, D. C. (2012). Compartment models of the diffusion MR signal in brain white matter: a taxonomy and comparison. NeuroImage, 59(3):2241–2254.

Perucho, J. A. U., Wang, M., Vardhanabhuti, V., Tse, K. Y., Chan, K. K. L., and Lee, E. Y. P. (2021). Association between IVIM parameters and treatment response in locally advanced squamous cell cervical cancer treated by chemoradiotherapy. European Radiology, pages 1–10. Publisher: Springer.

Pries, A. R. and Secomb, T. W. (2008). Blood flow in microvascular networks. In Micro-circulation, pages 3–36. Elsevier.

Rafael-Patino, J., Romascano, D., Ramirez-Manzanares, A., Canales-Rodríguez, E. J., Girard, G., and Thiran, J.-P. (2020). Robust monte-carlo simulations in diffusion-MRI: Effect of the substrate complexity and parameter choice on the reproducibility of results. Frontiers in neuroinformatics, 14:8. Publisher: Frontiers Media SA.

Salvaire, F. (2023). PySpice.

Schmid, F., Reichold, J., Weber, B., and Jenny, P. (2015). The impact of capillary dilation on the distribution of red blood cells in artificial networks. American Journal of Physiology-Heart and Circulatory Physiology, 308(7):H733–H742.

Scott, L. A., Dickie, B. R., Rawson, S. D., Coutts, G., Burnett, T. L., Allan, S. M., Parker, G. J., and Parkes, L. M. (2021). Characterisation of microvessel blood velocity and segment length in the brain using multi-diffusion-time diffusion-weighted mri. Journal of Cerebral Blood Flow & Metabolism, 41(8):1939–1953.

Stabinska, J., Zöllner, H. J., Thiel, T. A., Wittsack, H.-J., and Ljimani, A. (2023). Image downsampling expedited adaptive least-squares (ideal) fitting improves intravoxel incoherent motion (ivim) analysis in the human kidney. Magnetic Resonance in Medicine, 89(3):1055–1067.

Van, V. P., Schmid, F., Spinner, G., Kozerke, S., and Federau, C. (2021). Simulation of intravoxel incoherent perfusion signal using a realistic capillary network of a mouse brain. NMR in biomedicine, 34(7):e4528.

Veraart, J., Novikov, D. S., Christiaens, D., Ades-Aron, B., Sijbers, J., and Fieremans, E. (2016). Denoising of diffusion MRI using random matrix theory. NeuroImage, 142:394–406.

Wang, X., Song, J., Zhou, S., Lu, Y., Lin, W., Koh, T. S., Hou, Z., and Yan, Z. (2021). A comparative study of methods for determining intravoxel incoherent motion parameters in cervix cancer. Cancer Imaging, 21:1–11.

Weine, J., McGrath, C., Dirix, P., Buoso, S., and Kozerke, S. (2024). CMRsim–a python package for cardiovascular MR simulations incorporating complex motion and flow. Magnetic Resonance in Medicine. Publisher: Wiley Online Library.

Wetscherek, A., Stieltjes, B., and Laun, F. B. (2015). Flow-compensated intravoxel incoherent motion diffusion imaging. Magnetic resonance in medicine, 74(2):410–419.

Wu, D. and Zhang, J. (2019). Evidence of the diffusion time dependence of intravoxel incoherent motion in the brain. Magnetic resonance in medicine, 82(6):2225–2235. Publisher: Wiley Online Library.

